# A powerful replicability analysis of genome-wide association studies

**DOI:** 10.1101/2023.09.04.23295018

**Authors:** Yan Li, Haochen Lei, Xiaoquan Wen, Hongyuan Cao

**Affiliations:** School of Mathematics, Jilin University, Changchun, Jilin 130012, China; Department of Statistics, Florida State University, Tallahassee, FL 32306, USA; Department of Biostatistics, University of Michigan, Ann Arbor, MI 48109, USA

## Abstract

Replicability is the cornerstone of modern scientific research. Reliable identifications of genotype-phenotype associations that are significant in multiple genome-wide association studies (GWASs) provide stronger evidence for the findings. Current replicability analysis relies on the independence assumption among single nucleotide polymorphisms (SNPs) and ignores the linkage disequilibrium (LD) structure. We show that such a strategy may produce either overly liberal or overly conservative results in practice. We develop an efficient method, ReAD, to detect replicable SNPs associated with the phenotype from two GWASs accounting for the LD structure. The local dependence structure of SNPs across two heterogeneous studies is captured by a four-state hidden Markov model (HMM) built on two sequences of *p*-values. By incorporating information from adjacent locations via the HMM, our approach provides more accurate SNP significance rankings. ReAD is scalable, platform independent and more powerful than existing replicability analysis methods with effective false discovery rate (FDR) control. Through analysis of datasets from two asthma GWASs and two ulcerative colitis GWASs, we show that ReAD can identify replicable genetic loci that existing methods might otherwise miss.

## 1 Introduction

Genome-wide association studies (GWASs) allow for simultaneous study of millions of single nucleotide polymorphisms (SNPs). Numerous genetic risk variants associated with various phenotypes and complex diseases have been reported over the past couple of decades (McCarthy et al., 2008; MacArthur et al., 2017). These associations provide insights into the architecture of disease susceptibility. Despite these progresses, many reported genotype-phenotype associations fail to replicate in other studies (Ioannidis et al., 2001; Chanock et al., 2007). An analysis of past studies indicates that the cumulative prevalence of irreplicable preclinical research (including GWAS) exceeds 50% (Ioannidis, 2005; Prinz et al., 2011; Begley and Ellis, 2012; Freedman et al., 2015). Approximately 28 billion annually is spent on preclinical research that is not replicable in the United States alone (Freedman et al., 2015). Irreplicable and/or inconsistent between-study associations might be spurious findings caused by confounding factors, such as population stratification, misclassification of phenotypes, genotyping errors, or technical biases, among others. Replicability is now considered a *sine qua non* for establishing credible genotype-phenotype associations in the era of GWAS (Moonesinghe et al., 2008; Huffman, 2018). We study conceptual replicability where consistent results are obtained using different processes and populations that target the same scientific question. For GWASs, replicability analysis aims to detect genetic risk loci that are significantly associated with the same phenotype across different studies (Heller and Yekutieli, 2014; Heller et al., 2014; Bogomolov and Heller, 2022). By eliminating genetic associations that can not be generalized across studies, replicability analysis provides stronger support for genuine scientific findings, avoids wasted resources, and improves efficiency of drug development. This helps the translation of bench discoveries to bedsided therapies.

In GWASs, millions of SNPs are tested simultaneously, requiring multiple testing adjustment. False discovery rate (FDR), defined as the expectation of the proportion of false discoveries over total discoveries, is a commonly used metric for type I error control (Benjamini and Hochberg, 1995). A central characteristic of GWAS data is the linkage disequilibrium (LD) among SNPs, with which alleles at nearby sites can co-occur on the same haplotype more often than by chance alone (Pritchard and Przeworski, 2001; Wall and Pritchard, 2003). As a result, it is common to observe that phenotype-associated SNPs form clusters and exhibit high correlations within clusters (Wei et al., 2009). An effective approach to account for the LD structure among SNPs is through the hidden Markov model (HMM) (Churchill, 1992). Existing GWAS literature (Sun and Cai, 2009; Wei et al., 2009) using HMM for a single study is not applicable to replicability analysis of multiple studies. Furthermore, their approaches cannot be generalized to more than one study due to the heterogeneity of LD across different studies (Lonjou et al., 2003). Replicability analysis of GWASs explicitly accounting for the LD structure has not been studied before to the best of our knowledge.

To claim replicability, an *ad hoc* approach is to implement an FDR control method, such as the Benjamini and Hochberg (BH) procedure (Benjamini and Hochberg, 1995), for each study and intersect significant results from all studies as replicable findings. This approach does not control the FDR and moreover has low power as it does not borrow information from different studies. The maximum of *p*-values across studies (*P*_max_) is a straightforward significance measure for replicability (Benjamini et al., 2009). After summarizing data from multiple studies by *P*_max_, classic FDR control procedures such as BH are used for replicability analysis. This procedure is overly conservative as it guards against the worst scenario and does not incorporate the composite null structure of replicability analysis. For independent features from high-throughput experiments, various methods were proposed for replicability analysis. These methods are not robust to heterogeneity of different studies (Li et al., 2011; Philtron et al., 2018), require tuning parameters (Zhao et al., 2020), impose parametric assumptions on the *p*-values (Heller et al., 2014) or demand access to full datasets which can be prohibitive due to privacy concerns or logistics (McGuire et al., 2021).

We address the limitations of existing methods by developing an efficient method, ReAD (<u>Re</u>plicability <u>A</u>nalysis accounting for <u>D</u>ependence) to detect replicable genotype-phenotype associations across two GWASs by incorporating the LD structure. We use GWAS summary statistics such as *p*-values, treating multiple studies symmetrically. Our approach models the clustered signals from two studies with a four-dimensional HMM accounting for the heterogeneity of LD structures in different studies. Conditional on the HMM, we model the two *p*-value sequences as a four-group mixture of SNPs (Efron, 2012; Chung et al., 2014). The replicability null hypothesis consists of three components: zero effects in both studies, zero effect in one study and non-zero effect in another study and vice versa. ReAD calculates the posterior probability of replicability null given data. Compared to other replicability analysis methods, ReAD is robust as it is non-parametric, jointly models the signal and non-signal from different studies, and accounts for the heterogeneity of different studies. ReAD provides more efficient rankings of importance for replicable SNPs by pooling information from two *p*-value sequences via the forward and backward probabilities (Rabiner and Juang, 1986; Murphy, 2012). ReAD applies a step-up procedure to identify clusters of genotype-phenotype associated signals, improving the power of replicability analysis while effectively controlling the FDR. ReAD is computationally scalable to whole genome with tens of millions of SNPs. Its implementation combines the non-parametric expectation-maximization (EM) algorithm (Dempster et al., 1977) and the pool-adjacent-violator algorithm (PAVA) in shape constraint inference (Robertson et al., 1988; Busing, 2022), without any tuning parameters. We conduct extensive simulation studies to evaluate the performance of our approach across a wide range of scenarios. By applying our procedure to summary statistics of two asthma GWASs and two ulcerative colitis GWASs, we show that ReAD identifies more replicable genetic loci that otherwise might be missed using existing methods that do not account for the LD structure. These identified association signals pinpoint potential new loci on metabolisms and immunity.

## 2 Results

### 2.1 Method overview

ReAD takes *p*-values from two independent GWASs with the same phenotype as input. Suppose we have *J* SNPs with corresponding *p*-values (*p*_1*j*_, *p*_2*j*_), *j* = 1, …, *J*. We aim to identify replicable SNPs associated with the phenotype in both studies. Our method can handle SNPs in the whole genome where *J* is in the order of millions. We use *θ*_*ij*_ to represent the inferred association status of SNP *j* in study *i*. For each SNP, we consider its association analysis results are replicable if its corresponding *θ* values are consistently 1. The correlations between *θ*’s within a study are caused by LD among tested SNPs, and we model their dependence structure using a Markov chain. Following Li and Stephens (2003), this is an effective way to model the correlations between observed *p*-values. Given the observed *p*-values are from both studies, the overall model structure can be represented by a HMM.

We present our schematic in Figure 1. We use *s*_*j*_ *∈ {*0, 1, 2, 3*}* to denote the joint inferred association status for SNP *j*, where *s*_*j*_ = 0 if *θ*_1*j*_ = *θ*_2*j*_ = 0, *s*_*j*_ = 1 if *θ*_1*j*_ = 0 and *θ*_2*j*_ = 1, *s*_*j*_ = 2 if *θ*_1*j*_ = 1 and *θ*_2*j*_ = 0, and *s*_*j*_ = 3 if *θ*_1*j*_ = *θ*_2*j*_ = 1. The composite null for replicability analysis corresponds to *s*_*j*_ *∈ {*0, 1, 2*}*. To capture the local dependence of LD structure among SNPs, we impose a four-state HMM on ***s*** = (*s*_1_, …, *s*_*J*_). The transition matrix is denoted as *A* = *{a*_*kl*_ : *k, l* = 0, 1, 2, 3*}*, where the transition probability from *s*_*j*_ = *k* to *s*_*j*+1_ = *l* is given by *a*_*kl*_, and 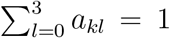 for all *k*. An efficient EM algorithm in combination with the forward-backward procedure and PAVA is developed to estimate the unknown parameters and functions. We use the posterior probability of being replicability null, rLIS_*j*_, *j* = 1, …, *J*, as the test statistic and obtain 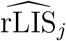 for all SNPs. By applying a step-up procedure on 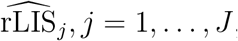, we get powerful testing results while controlling the FDR. More details of ReAD can be found in the Methods Section and the Supplemental Note A.

**Figure 1:**
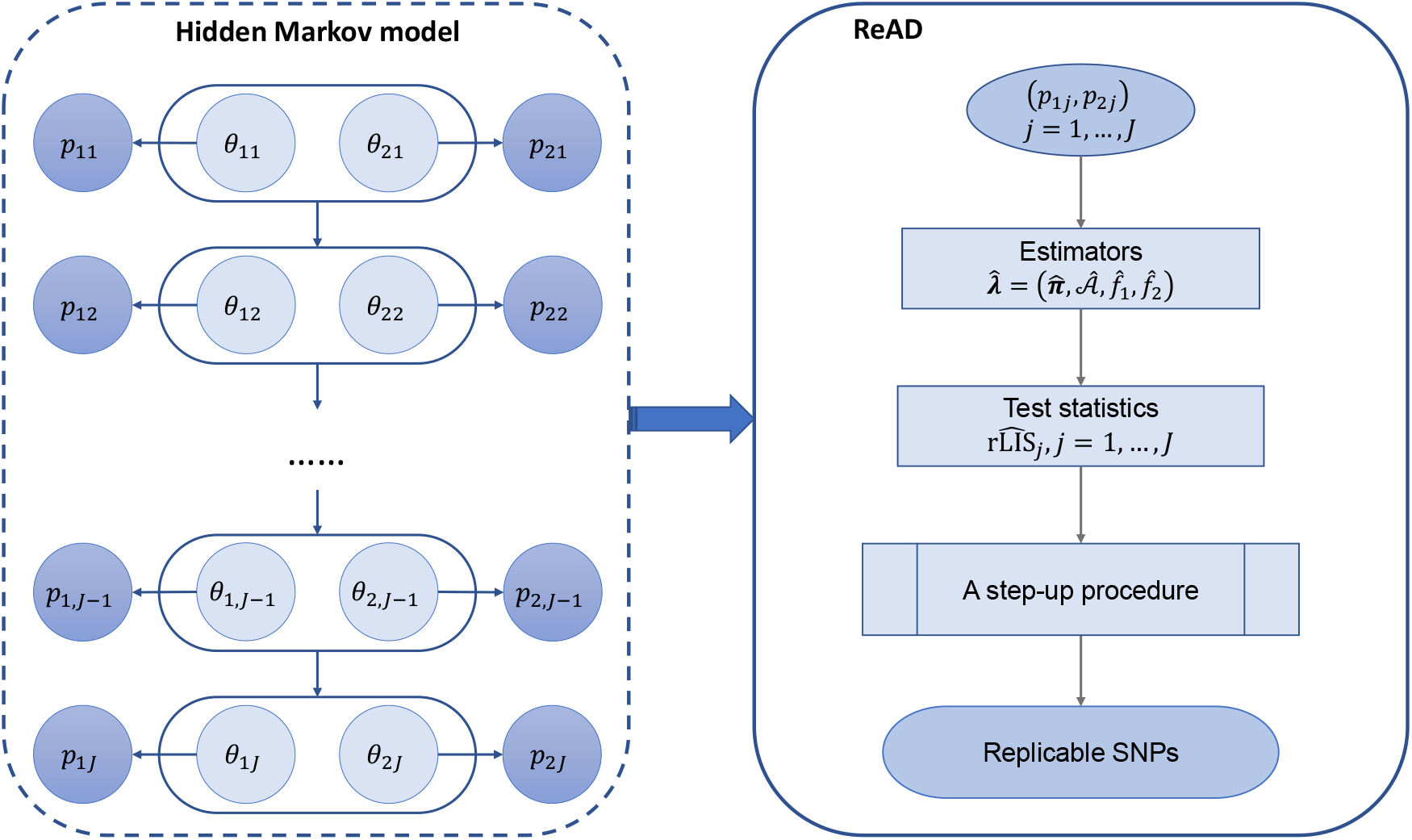
Schematic of ReAD. *θ*_*ij*_ represents the inferred association status of SNP *j* (*j* = 1, …, *J*) in study *i* (*i* = 1, 2). For each SNP *j*, we consider its association analysis results are replicable if *θ*_1*j*_ = *θ*_2*j*_ = 1. The dependence structure among SNPs across two studies can be modeled with a HMM.

### 2.2 Simulation study

#### 2.2.1 Simulation I

In simulation I, we evaluated the FDR and statistical power of ReAD based on the rLIS statistic across two studies. Here power is defined as the averaged proportion of true discoveries among the total number of non-null hypotheses. We compare the FDR and power of ReAD with several replicability analysis methods developed under independence, including the *ad hoc* BH method, the MaxP method based on *P*_max_(Benjamini et al., 2009) and the STAREG method based on the local false discovery rate (Lfdr) (Li et al., 2023). Details of these methods can be found in the Supplemental Note B. An extensive comparison with more replicability analysis methods can be found in Supplemental Note C.

In each simulation, the hidden states of 10, 000 SNPs were generated from a four-state Markov chain. A detailed description of the data generating process is provided in Supplemental Note C. In all simulations, we fix the initial distribution of four states as ***π*** = (0.9, 0.025, 0.025, 0.05). The signals from two studies are generated from normal distributions with mean *µ*_*i*_ and variance 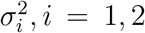. We vary the transition matrix *A* = *{a*_*kl*_ : *k, l* = 0, 1, 2, 3*}* and *µ*_2_ while fix *µ*_1_ = 2, and *σ*_1_ = *σ*_2_ = 1. Empirical FDR and power are calculated from 100 replications for each setting. The results are summarized in Figure 2 (left: FDR; right: power). In Figure 2, each row corresponds to a different *a*_00_, and each column corresponds to a different *a*_33_. In each panel, we set *µ*_2_ to 1.5, 2, or 3. At FDR level 0.05, we see that the *ad hoc* BH fails to control the FDR. MaxP is overly conservative across all settings.

**Figure 2:**
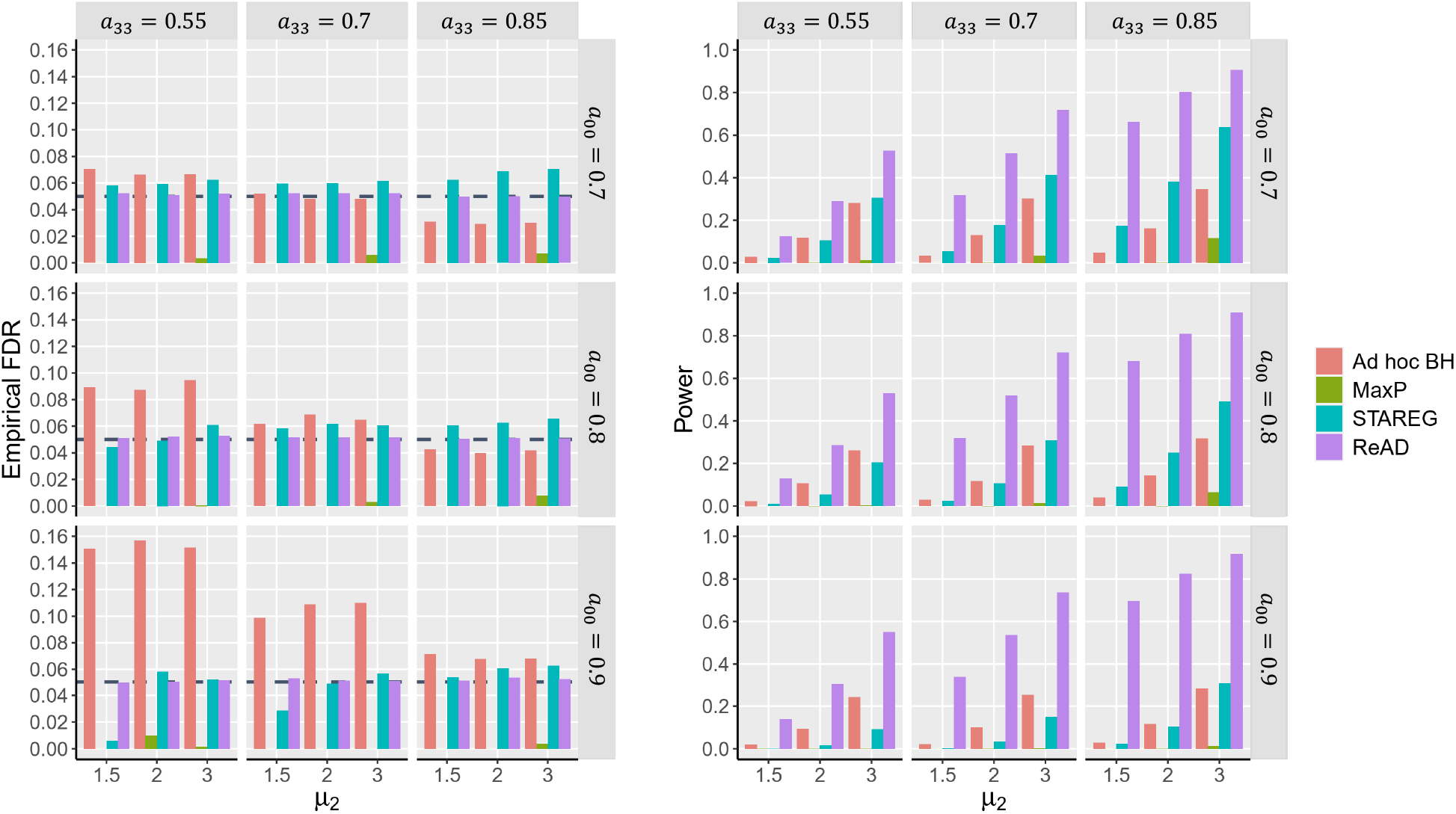
FDR control and power comparison of different methods.

STAREG has a slight FDR inflation in some settings. By accounting for the local dependence structure via the rLIS statistic, ReAD properly controls the FDR and has substantial power gain compared to competing methods. The powers of all methods increase as *µ*_2_ increases.

The forward-backward procedure of HMM implies that a small rLIS does not occur alone, but in clusters. Therefore, ReAD tends to identify the entire cluster of genotype-phenotype associations. Such clusters are unlikely to occur by chance and are more plausible biological signals. To illustrate this, *p*-values for two studies are generated following the above strategy by setting *a*_00_ = 0.9, *a*_33_ = 0.7, and *µ*_2_ = 2. We compare three methods for testing the composite replicability null hypotheses across two studies: the MaxP method (Benjamini et al., 2009), the STAREG method (Li et al., 2023), and the ReAD method. Figure 3 presents results of different methods in one replication. It can be seen that MaxP is extremely conservative, which only identifies one single signal; STAREG rejects individual hypotheses with very small *p*-values in both studies; whereas ReAD can identify clusters of replicable signals.

**Figure 3:**
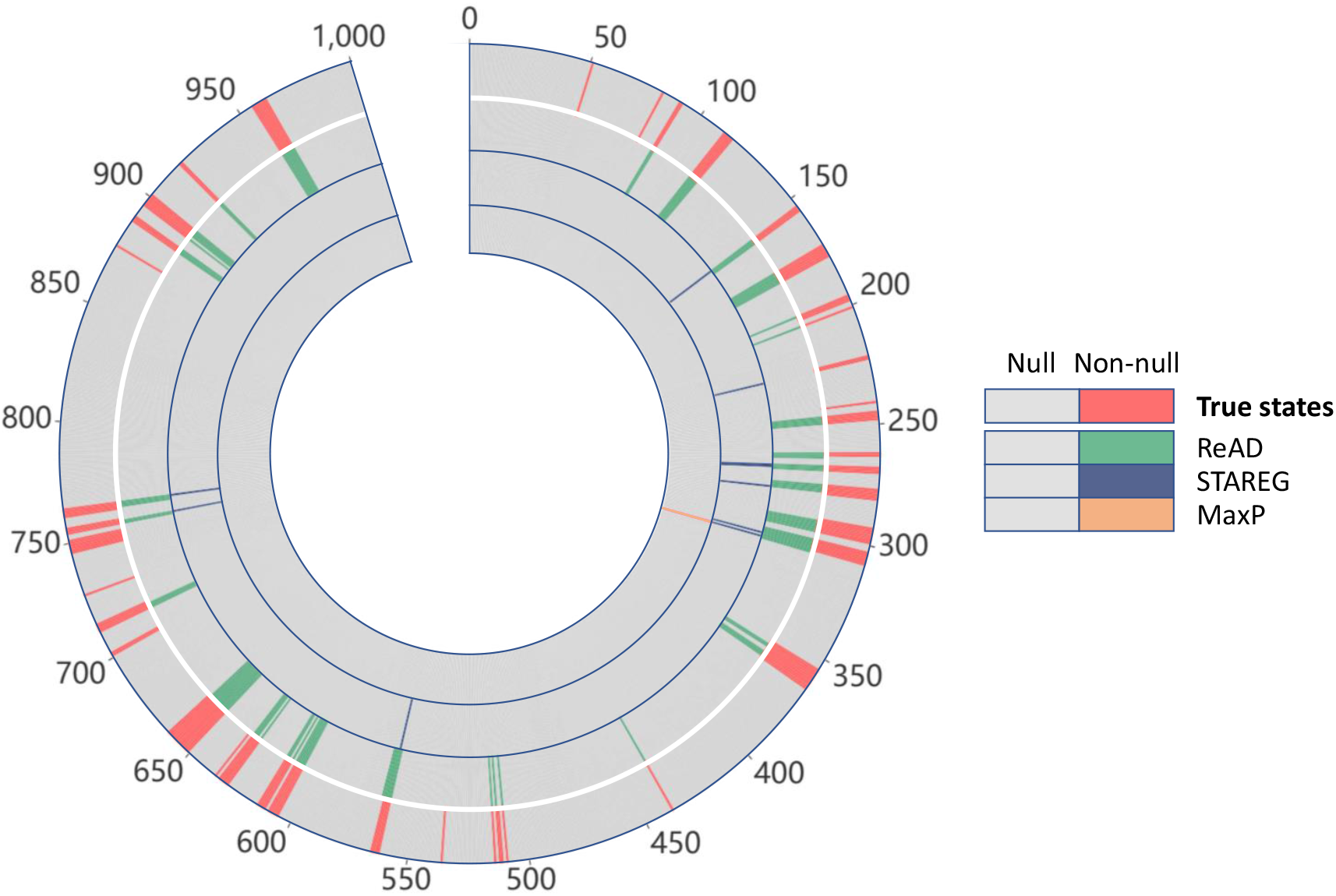
Methods comparison for cluster identification. Circles range from 1 (the outermost circle) to 4 (the innermost circle). The outermost circle represents true states; circle 2 represents ReAD, circle 3 represents STAREG and cirle 4 represents MaxP.

#### 2.2.2 Simulation II

By incorporating the LD structure in GWASs through HMM, the rLIS statistic integrates information from adjacent locations. Therefore, the rankings of SNPs based on rLIS are different from the rankings from MaxP (based on *P*_max_) and STAREG (based on Lfdr). We perform simulation studies to demonstrate different rankings in GWASs with realistic LD patterns among SNPs. Data for two studies are generated based on two SNP matrices from the Genetic European Variation in Disease project (Lappalainen et al., 2013). The CEU genotype data are collected from 78 Utah residents with Northern and Western European ancestry, and the FIN genotype data are measured from 89 Finnish in Finland. CEU and FIN are both sub-populations of the European Ancestry population, therefore they may have similar LD structures. A detailed description of the strategy to generate continuous phenotypes corresponding to SNPs in a single gene across two studies is provided in the Methods Section. To make realistic simulations, we adjust the signal-to-noise ratio in two studies so that the SNP heritability 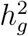 is centered between 0.2 and 0.3. Figure 4(A) presents the histogram of 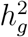 for CEU and FIN studies generated from 5, 000 replications. We repeat the above data generating process to simulate GWAS data for 100 genes, resulting in 1, 676, 400 pairs of *p*-values for corresponding SNPs. As in Wei et al. (2009), we define five adjacent SNPs on each side of the 400 causal SNPs as relevant SNPs and evaluate the performance of different replicability analysis methods by calculating the percentages of selecting relevant SNPs.

**Figure 4:**
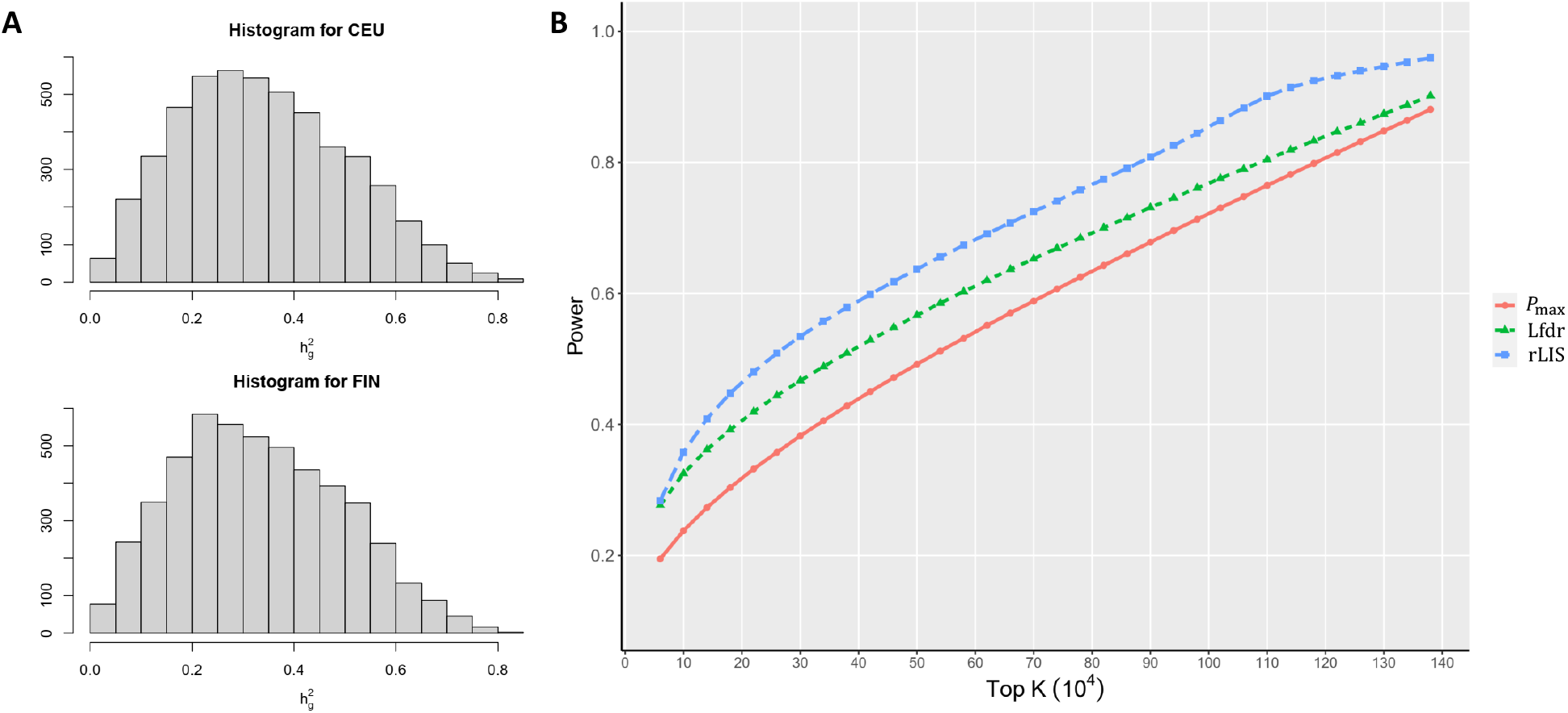
(A) Histogram of estimated 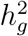 for simulated CEU data and FIN data from 5, 000 runs. (B) Power of top *K* SNPs identified by different methods. The power is calculated as the averaged percentages of true positives selected by the top *K* SNPs ranked based on rLIS, Lfdr produced by STAREG, and *P*_max_ used in MaxP. The results are calculated from 100 runs.

We average the percentages of true positives selected by the top *K* hits from 100 runs as our evaluation criterion. The power curves of different *K* values based on SNP rankings produced by *P*_max_, Lfdr and rLIS are depicted in Figure 4(B). We see that rLIS shows higher power than *P*_max_ and Lfdr, indicating that the rankings based on rLIS are more efficient than the rankings based on *P*_max_ and Lfdr in replicability analysis of GWAS data by incorporating the LD block structure through HMM.

### 2.3 Data analysis

#### 2.3.1 Replicability analysis of asthma GWASs

Asthma is a complex bronchial disease characterized by chronic inflammation and narrowing of the airways, which is caused by a combination of environmental and genetic factors. The prevalence of asthma varies across different populations and ethnicities. We implement ReAD to conduct replicability analysis of asthma GWASs from the Trans-National Asthma Genetic Consortium (TAGC) and UK Biobank. The results are compared with competing methods. Demenais et al. (2018) conducted ancestry-specific meta-analyses from ethnically-diverse populations and deposited the HapMap2-imputed data in the TAGC consortium. The TAGC asthma GWAS data with high-density genotyped and imputed SNP based on the European-ancestry comprises 8, 843, 303 genetic variants for 19, 954 asthma cases and 107, 715 controls. UK Biobank is a large-scale prospective cohort study with over half a million participants aged 40-69 years from the United Kingdom between 2006 and 2010 (Sudlow et al., 2015). The imputed asthma GWAS from UK Biobank contains summary statistics for 8, 856, 162 genetic variants measured on 39, 049 self-reported asthma cases and 298, 070 controls. We filter out SNPS with minor allele frequency (MAF) smaller than 0.05, resulting in 6, 234, 241 SNPs in the TAGC study and 6, 242, 120 SNPs in UK Biobank. After taking the intersection of SNPs in the two studies, we obtain paired *p*-values of 6, 222, 195 SNPs to conduct replicability analysis.

As the *ad hoc* BH does not control FDR, we apply MaxP and STAREG on the paired *p*-values for comparison. The GWAS Catalog (Welter et al., 2014) reported cytogenetic regions (loci) associated with asthma. To assess the replicability of GWAS loci, we state that if at least one of the identified SNPs falls into one of the regions, the locus is identified as replicable. If a locus contains multiple significant SNPs, the SNP with the strongest association is considered as the lead SNP. For instance, if we use STAREG with Lfdr as the test statistic, the SNP with the smallest Lfdr is the lead SNP.

At FDR level 5 *×* 10^*−*8^, MaxP identifies 2, 853 significant SNPs in 10 loci, which are also identified by STAREG and ReAD. Compared to MaxP, STAREG identifies 909 additional significant SNPs in 3 loci. By capturing the local LD structure through HMM, ReAD identifies 10, 084 significant SNPs in 28 genetic loci with replicable asthma associations, of which 15 loci are not detected by MaxP or STAREG. Figure 5 presents the Manhattan plots of MaxP, STAREG, and ReAD. In Figure 5, the vertical axis are *−* log_10_ transformations of test statistics for replicability analysis, i.e., *P*_max_ for MaxP, Lfdr for STAREG, and rLIS for ReAD.

**Figure 5:**
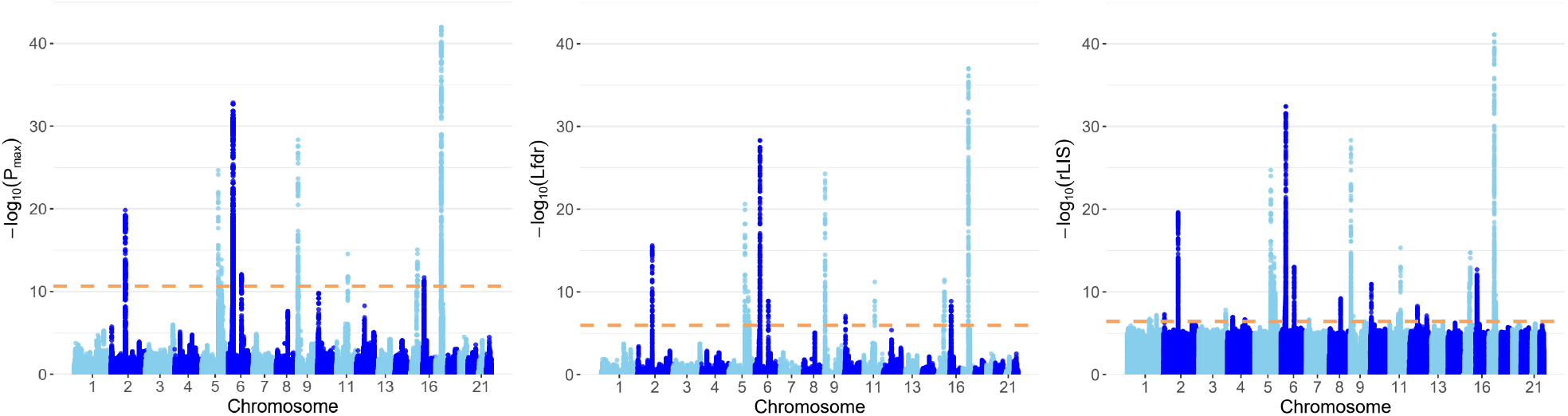
The Manhattan plots based on *P*_max_, Lfdr and rLIS. The dashed horizontal lines denote the FDR cutoffs of 5 *×* 10^*−*8^ produced by MaxP, STAREG, and ReAD, respectively.

Table 1 displays main characteristics of the 28 cytogenetic regions identified by ReAD. The mapped gene denotes genes overlapping or closest to the lead SNP in the identified locus. The 15 loci only identified by ReAD harbor signals closely related to asthma. For example, the lead SNP in locus 2p25.1, rs10174949, is in the intron of gene *LINC00299* and plays an important role in atopic dermatitis, including asthma, hay fever and eczema in European and UK populations (Zhu et al., 2020, 2018; Ferreira et al., 2017). The 8q21.13 region is reported to be associated with asthma and hay fever in a European-ancestry study (Ferreira et al., 2014). The lead SNP rs6473226 lies between gene *MIR5708* (chr8:80,241,389–80,241,473) and gene *RNU6-1213P* (chr8:80,405,516–80,405,609), and its association with asthma has been observed in several European-ancestry studies (Demenais et al., 2018; Olafsdottir et al., 2020).

**Table 1:**
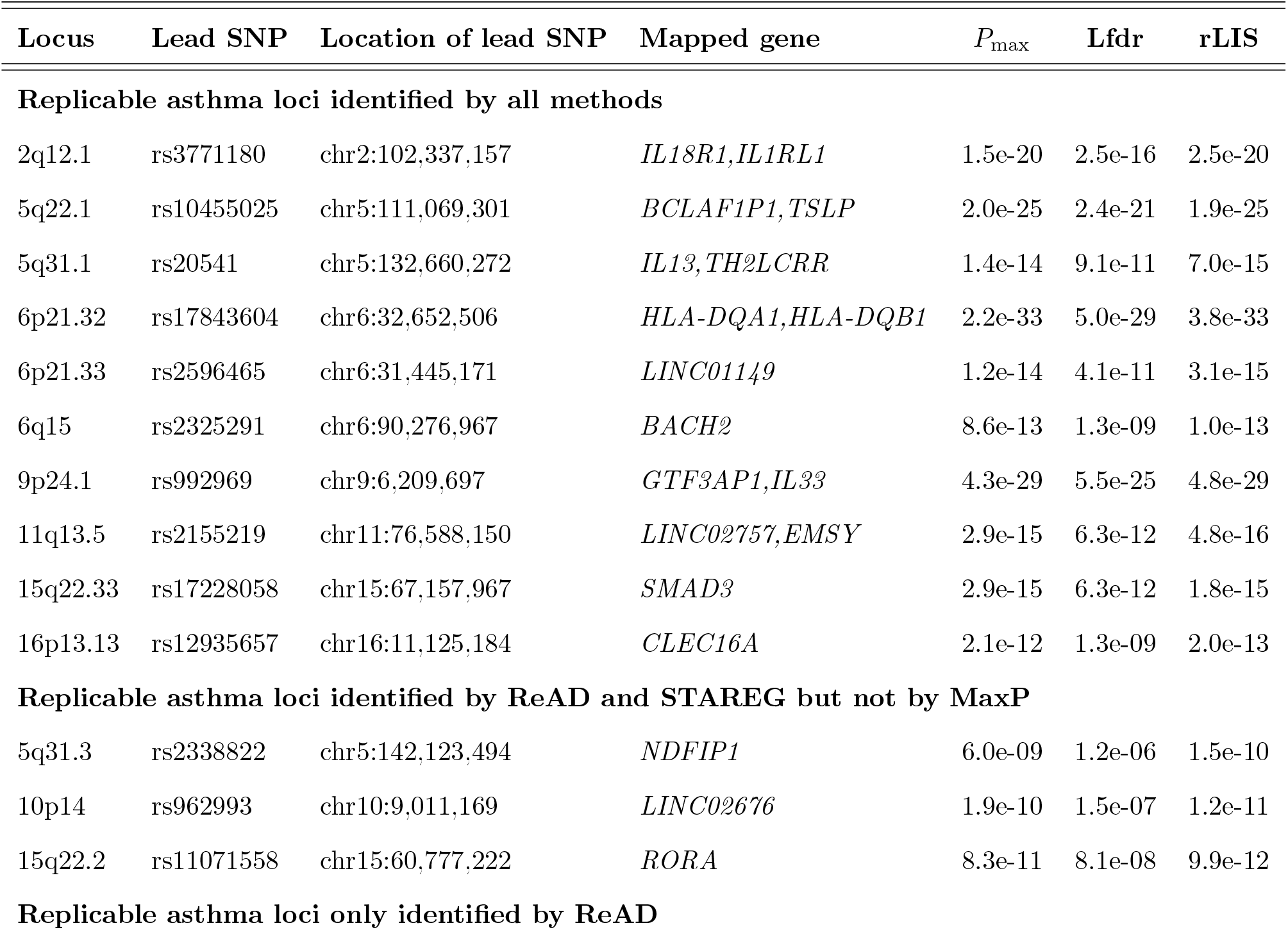
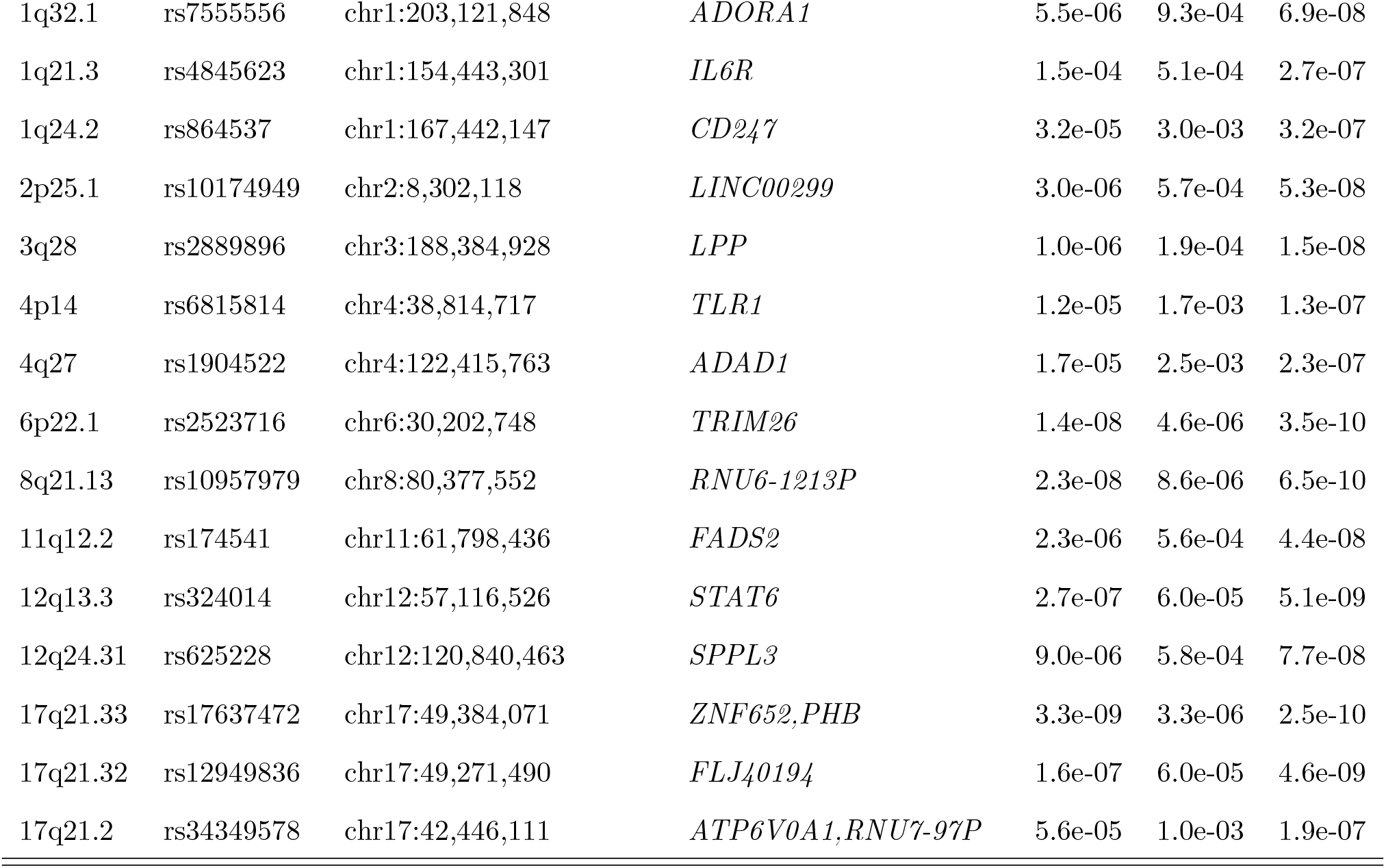
Main characteristics of the 28 loci associated with asthma in the European-ancestry TAGC and UK Biobank GWASs identified by ReAD. The SNP with the strongest association within each locus is called Lead SNP. The mapped gene denotes genes overlapping or closest to the lead SNP in the identified locus.

| Locus | Lead SNP | Location of lead SNP | Mapped gene | $P_{\max}$ | Lfdr | rLIS |
| --- | --- | --- | --- | --- | --- | --- |
| **Replicable asthma loci identified by all methods** |  |  |  |  |  |  |
| 2q12.1 | rs3771180 | chr2:102,337,157 | *IL18R1,IL1RL1* | 1.5e-20 | 2.5e-16 | 2.5e-20 |
| 5q22.1 | rs10455025 | chr5:111,069,301 | *BCLAF1P1,TSLP* | 2.0e-25 | 2.4e-21 | 1.9e-25 |
| 5q31.1 | rs20541 | chr5:132,660,272 | *IL13,TH2LCRR* | 1.4e-14 | 9.1e-11 | 7.0e-15 |
| 6p21.32 | rs17843604 | chr6:32,652,506 | *HLA-DQA1,HLA-DQB1* | 2.2e-33 | 5.0e-29 | 3.8e-33 |
| 6p21.33 | rs2596465 | chr6:31,445,171 | *LINC01149* | 1.2e-14 | 4.1e-11 | 3.1e-15 |
| 6q15 | rs2325291 | chr6:90,276,967 | *BACH2* | 8.6e-13 | 1.3e-09 | 1.0e-13 |
| 9p24.1 | rs992969 | chr9:6,209,697 | *GTF3AP1,IL33* | 4.3e-29 | 5.5e-25 | 4.8e-29 |
| 11q13.5 | rs2155219 | chr11:76,588,150 | *LINC02757,EMSY* | 2.9e-15 | 6.3e-12 | 4.8e-16 |
| 15q22.33 | rs17228058 | chr15:67,157,967 | *SMAD3* | 2.9e-15 | 6.3e-12 | 1.8e-15 |
| 16p13.13 | rs12935657 | chr16:11,125,184 | *CLEC16A* | 2.1e-12 | 1.3e-09 | 2.0e-13 |
| **Replicable asthma loci identified by ReAD and STAREG but not by MaxP** |  |  |  |  |  |  |
| 5q31.3 | rs2338822 | chr5:142,123,494 | *NDFIP1* | 6.0e-09 | 1.2e-06 | 1.5e-10 |
| 10p14 | rs962993 | chr10:9,011,169 | *LINC02676* | 1.9e-10 | 1.5e-07 | 1.2e-11 |
| 15q22.2 | rs11071558 | chr15:60,777,222 | *RORA* | 8.3e-11 | 8.1e-08 | 9.9e-12 |
| **Replicable asthma loci only identified by ReAD** |  |  |  |  |  |  |
| 1q32.1 | rs7555556 | chr1:203,121,848 | *ADORA1* | 5.5e-06 | 9.3e-04 | 6.9e-08 |
| 1q21.3 | rs4845623 | chr1:154,443,301 | *IL6R* | 1.5e-04 | 5.1e-04 | 2.7e-07 |
| 1q24.2 | rs864537 | chr1:167,442,147 | *CD247* | 3.2e-05 | 3.0e-03 | 3.2e-07 |
| 2p25.1 | rs10174949 | chr2:8,302,118 | *LINC00299* | 3.0e-06 | 5.7e-04 | 5.3e-08 |
| 3q28 | rs2889896 | chr3:188,384,928 | *LPP* | 1.0e-06 | 1.9e-04 | 1.5e-08 |
| 4p14 | rs6815814 | chr4:38,814,717 | *TLR1* | 1.2e-05 | 1.7e-03 | 1.3e-07 |
| 4q27 | rs1904522 | chr4:122,415,763 | *ADAD1* | 1.7e-05 | 2.5e-03 | 2.3e-07 |
| 6p22.1 | rs2523716 | chr6:30,202,748 | *TRIM26* | 1.4e-08 | 4.6e-06 | 3.5e-10 |
| 8q21.13 | rs10957979 | chr8:80,377,552 | *RNU6-1213P* | 2.3e-08 | 8.6e-06 | 6.5e-10 |
| 11q12.2 | rs174541 | chr11:61,798,436 | *FADS2* | 2.3e-06 | 5.6e-04 | 4.4e-08 |
| 12q13.3 | rs324014 | chr12:57,116,526 | *STAT6* | 2.7e-07 | 6.0e-05 | 5.1e-09 |
| 12q24.31 | rs625228 | chr12:120,840,463 | *SPPL3* | 9.0e-06 | 5.8e-04 | 7.7e-08 |
| 17q21.33 | rs17637472 | chr17:49,384,071 | *ZNF652,PHB* | 3.3e-09 | 3.3e-06 | 2.5e-10 |
| 17q21.32 | rs12949836 | chr17:49,271,490 | *FLJ40194* | 1.6e-07 | 6.0e-05 | 4.6e-09 |
| 17q21.2 | rs34349578 | chr17:42,446,111 | *ATP6V0A1,RNU7-97P* | 5.6e-05 | 1.0e-03 | 1.9e-07 |

#### 2.3.2 Replicability analysis of ulcerative colitis GWASs

Inflammatory bowel disease is a chronic, relapsing intestinal inflammatory disease. It has the highest age-standardized prevalence rate in the US followed by the UK (Alatab et al., 2020) with increasing prevalence in Asia and developing countries (Molodecky et al., 2012). Ulcerative colitis (UC) is one of the two main forms of inflammatory bowel disease. We conduct replicability analysis of GWASs from the International Inflammatory Bowel Disease Genetics Consortium (IIBDGC) and the UK Biobank. The IIBDGC GWAS analyses 8, 857, 076 SNPs from 6, 968 UC cases and 20, 464 population controls of European descent (Liu et al., 2015). The imputed UK Biobank GWAS data contain summary statistics of 8, 856, 162 SNPs genotyped on 1, 795 self-reported UC cases and 335, 324 controls from the United Kingdom. We filter out SNPs with MAF smaller than 0.05, resulting in 6, 243, 744 SNPs in the IIBDGC study and 6, 242, 120 SNPs in the UK Biobank. We use the paired *p*-values of 6, 232, 147 SNPs common to both studies as input for replicability analysis.

We apply MaxP, STAREG, and ReAD on the paired *p*-values. At FDR level 5 *×* 10^*−*8^, MaxP identifies 1, 239 significant SNPs in 1 locus. STAREG identifies 1, 542 significant SNPs in 2 loci, one of which is also detected by MaxP. ReAD identifies 3, 307 significant SNPs in 7 genetic loci, including 5 loci that are not detected by MaxP or STAREG. Figure 6 presents the Manhattan plots of MaxP, STAREG and ReAD.

**Figure 6:**
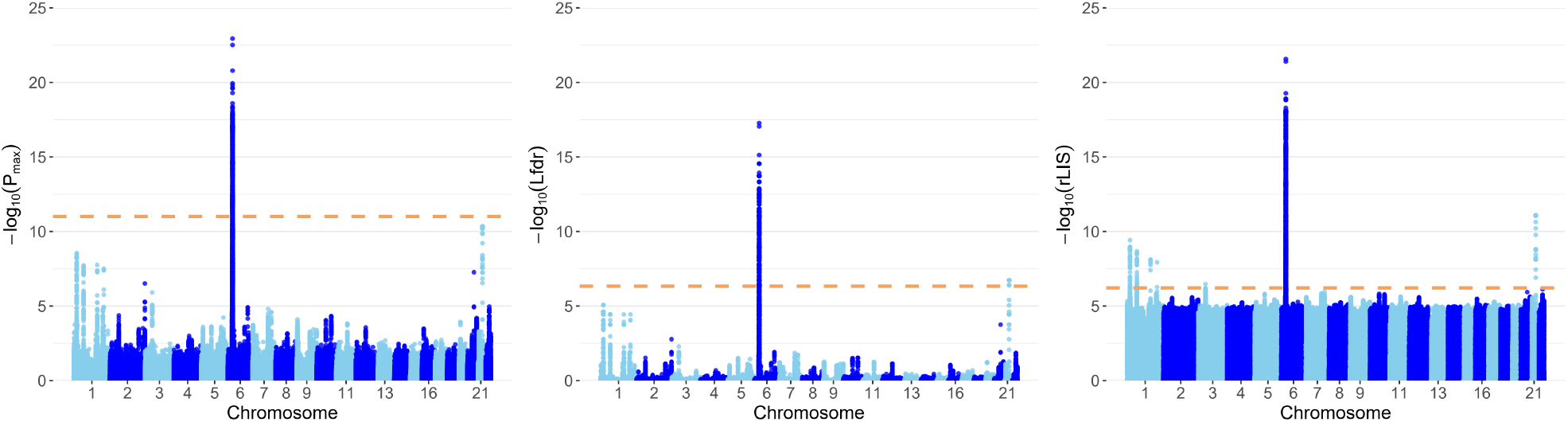
The Manhattan plots based on *P*_max_, Lfdr and rLIS. The dashed horizontal lines denote the FDR cutoffs of 5 *×* 10^*−*8^ produced by MaxP, STAREG, and ReAD, respectively.

We assess the replicability of genetic loci identified by different methods in GWAS Catalog (Welter et al., 2014). Table 2 presents the main characteristics of the 7 replicable genetic loci identified by ReAD. UC associations of these loci in cohorts of European descent have been repored in the literature. For instance, the lead SNP of loci 6p21.32, rs6927022, is in the intron of gene *HLA-DQA1*, and the HLA complex is associated with multiple risk alleles for inflammatory bowel disease, including UC (Nowak et al., 2021; Reinshagen et al., 1996; Ashton et al., 2019). The lead SNP harbored in loci 1q23.3, rs1801274, is only identified by ReAD, and has confirmed associations with UC in several European-ancestry studies (Liu et al., 2015; De Lange et al., 2017; Anderson et al., 2011). We have additional validations in DisGeNET, a versatile platform that contains a comprehensive catalog of genes and variants associated with human diseases (Piñero et al., 2015). Many mapped genes of the lead SNP only identified by ReAD have been reported to be associated with UC, such as gene *FCGR2A* in locus 1q23.3, gene *IL23R* in locus 1p31.3, gene *IL10* in locus 1q32.1, and gene *MST1* in locus 3p21.31.

**Table 2:** Main characteristics of the 7 loci associated with UC in the European-ancestry IIB-DGC and UK Biobank GWASs identified by ReAD. The SNP with the strongest association within each locus is called the lead SNP. The mapped gene denotes genes overlapping or closest to the lead SNP in the identified locus.

| Locus | Lead SNP | Location of lead SNP | Mapped gene | $P_{\max}$ | Lfdr | rLIS |
| --- | --- | --- | --- | --- | --- | --- |
| **Replicable asthma loci identified by all methods** |  |  |  |  |  |  |
| 6p21.32 | rs6927022 | chr6:32,644,620 | *HLA-DQA1* | 1.1e-20 | 2.8e-15 | 1.2e-19 |
| **Replicable asthma loci identified by ReAD and STAREG but not by MaxP** |  |  |  |  |  |  |
| 21q22.2 | rs2836882 | chr21:39,094,644 | *RPL23AP12* | 4.5e-11 | 1.9e-07 | 8.1e-12 |
| **Replicable asthma loci only identified by ReAD** |  |  |  |  |  |  |
| 1p36.13 | rs4654903 | chr1:19,874,497 | *RNF186, OTUD3* | 1.3e-08 | 3.6e-05 | 6.7e-09 |
| 1q23.3 | rs1801274 | chr1:161,509,955 | *FCGR2A* | 1.7e-08 | 3.6e-05 | 7.7e-09 |
| 1p31.3 | rs2201841 | chr1:67,228,519 | *C1orf141, IL23R* | 6.1e-08 | 1.8e-04 | 1.8e-08 |
| 1q32.1 | rs3024505 | chr1:206,766,559 | *Y-RNA, IL10* | 4.1e-08 | 8.9e-05 | 5.4e-07 |
| 3p21.31 | rs3197999 | chr3:49,684,099 | *MST1* | 1.2e-06 | 5.2e-03 | 3.4e-07 |

## 3 Discussion

In this paper, we present ReAD, an efficient method accounting for the LD structure to identify replicable associations from two GWASs datasets. We conduct extensive simulation studies and analyze two GWAS datasets. Compared to conventional approaches that impose independence assumption among SNPs, ReAD provides effective FDR control. It has a substantial power gain in identifying genuine and replicable genetic loci. It is computationally scalable to hundreds of millions of SNPs and has no tuning parameters.

In this paper, our discussion mainly focuses on assessing the replicability of each SNP within a genomic locus. We acknowledge that, in the applications of genetic association analysis, varying LD patterns between studies can lead to inconsistent significant findings at the SNP level. Hence scientifically, a more relevant question should be the consistency of underlying association signals within each interrogated locus across original and replication studies. To this end, we apply a simple and practical strategy requiring at least one SNP-level findings replicable. With the potential varying LD structures fully accounted for by the proposed HMM, we find this strategy intuitive and effective when applied to genomic loci with proper resolutions (as illustrated by our simulations and real data examples). Nevertheless, this locus-level criterion may be considered overly lenient. We will continue to explore alternative locus-level replicability assessment criteria in our future work.

In this work, we use repeated significance to assess replicability. We note that applying such a replicability criterion is debatable in the scientific community. While acknowledging its drawbacks, especially its conservativeness, we note the following context-specific factors. First, despite continued efforts to include more informative statistics summarizing GWAS findings, a large body of historical GWAS findings are *only* reported in *p*-values (See GWAS catalog (Welter et al., 2014)), which fundamentally limits applying alternative replicability criteria. Second, because complicated unknown confoundings, e.g., population stratification and unobserved batch effects in genotyping experiments, often cause false positives in genetic association analysis, the genetics community has consistently advocated conservative replicability criteria to ensure the reliability of GWAS findings (Skol et al., 2006; McGuire et al., 2021). Third, we emphasize that our main statistical contribution is to account for the correlation structure between genetic variants, and our work can be naturally extended to applying other alternative replicability criteria.

On a related point, although we exclusively assume that GWAS results are reported in the form of single-SNP testing *p*-values throughout this paper, the proposed statistical methodology can be extended to other forms of summary statistics. For example, probabilistic fine-mapping analysis of genetic association signals has become increasingly popular, thanks to the availability of efficient variable selection algorithms (Benner et al., 2016; Wang et al., 2020; Wen et al., 2016). The fine-mapping result is typically given as a posterior inclusion probability (PIP) at the individual SNP level. With the ability to construct a Bayesian credible set for each underlying signal within a genomic locus, the PIPs have many advantages over single-SNP *p*-values. Theoretically, our work can be straightforwardly extended to this setting by noting the connection that 1 *−* PIP is equivalent to the local fdr in the Bayesian perspective. We will leave this extension to our future work.

## 4 Methods

### 4.1 The hidden Markov model for replicability analysis

Suppose there are *J* SNPs in two independent GWASs. We are interested in testing whether the *j*th SNP is associated with the phenotype in both studies. Let *θ*_*ij*_ denote the inferred association status of SNP *j* in study *i*, where *θ*_*ij*_ = 1 indicates the *j*th SNP (*j* = 1, …, *J*) is inferred associated with the phenotype in study *i* (*i* = 1, 2) and *θ*_*ij*_ = 0 otherwise. We use *s*_*j*_ (*j* = 1, …, *J*) to denote the joint status.

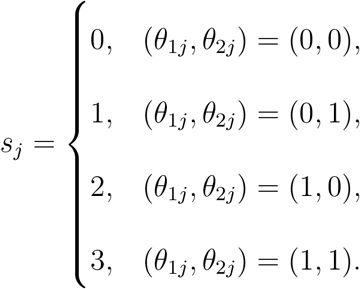

The replicability null hypotheses is

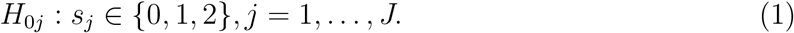

Let 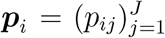 denote *p*-values of *J* SNPs in study *i*. We use mixture models for the conditional distributions of *p*-values given *θ* values. Specifically,

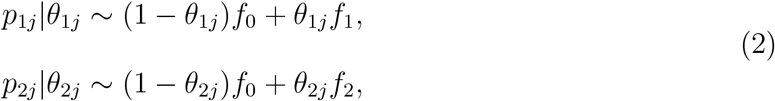

where *f*_0_ is the probability density function of *p*-values when *θ*_1*j*_ = *θ*_2*j*_ = 0, and *f*_1_ and *f*_2_ are the *p*-value density functions under non-null in study 1 and study 2, respectively. We assume *f*_0_ follows the standard uniform distribution and impose the following monotone likelihood ratio condition (Sun and Cai, 2007; Cao et al., 2013, 2022).

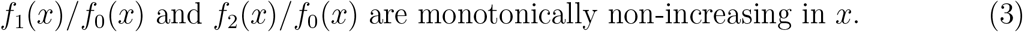

This condition naturally arises as small *p*-values indicate evidence against the null. To capture the LD structure among SNPs, we assume that ***s*** = (*s*_1_, …, *s*_*J*_) follows a four-state stationary, irreducible, and aperiodic hidden Markov model (HMM). The transition probabilities

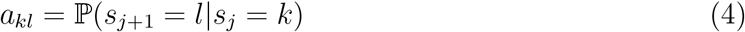

for *k, l* = 0, 1, 2, 3 with constraint 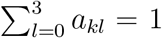. The stationary distribution of each state *s*_*j*_ is 𝕡 (*s*_*j*_ = *k*) = *π*_*k*_ for *k* = 0, 1, 2, 3 and 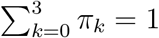. The paired *p*-values for the *j*th SNP are assumed to be conditionally independent satisfying

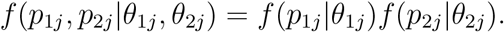

Based on the mixture model (2), we have

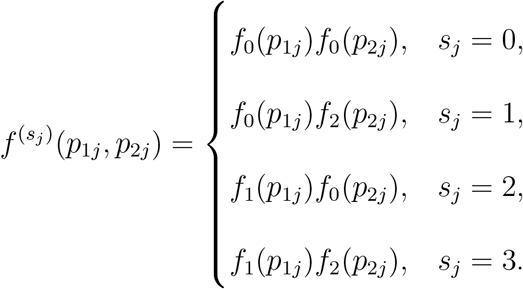

Denote by *𝒜* = *{a*_*kl*_ : *k, l* = 0, 1, 2, 3*}* the transition matrix, ***π*** = (*π*_0_, *π*_1_, *π*_2_, *π*_3_) the vector of stationary distribution, and *ℱ* = *f* ^(0)^, *f* ^(1)^, *f* ^(2)^, *f* ^(3)^ the probability density functions of the bivariate observations (*p*_1*j*_, *p*_2*j*_). The convergence theorem of a Markov chain (Theorem 5.5.1 in Durrett (2019)) implies that

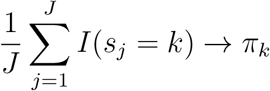

almost surely for *k* = 0, 1, 2, 3 as *J → ∞*. As *f*_0_ is assumed to follow a standard uniform distribution, we use ***λ*** = (***π***, *𝒜, f*_1_, *f*_2_) to denote the collection of unknown parameters and functions in the HMM. Our goal is to separate the replicable SNPs (*s*_*j*_ = 3) from the non-replicable SNPs (*s*_*j*_ *∈ {*0, 1, 2*}*) based on the observed bivariate *p*-values.

### 4.2 FDR control for replicability analysis accounting for LD

#### 4.2.1 The rLIS statistic for replicability analysis across two studies

Consider the ideal setup that an oracle knows ***λ*** = (***π***, *𝒜, f*_1_, *f*_2_). We define the replicability local index of significance (rLIS) as the posterior probability of being null. Specifically,

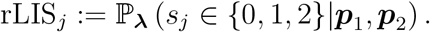

Given ***λ***, the forward and backward probabilities are defined as 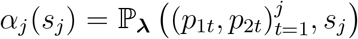, and 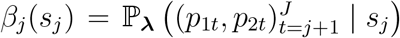, respectively. The forward-backward procedure (Baum et al., 1970) can be used in the calculation. Specifically, we initialize 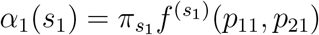 and *β*_*J*_ (*s*_*J*_) = 1. We can obtain *α*_*j*_(*·*) and *β*_*j*_(*·*) for *j* = 1, …, *J* recursively by

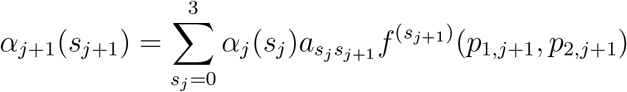

and

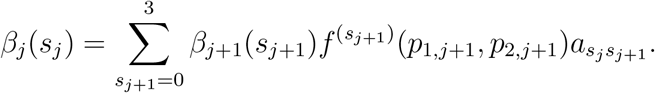

Hence we have

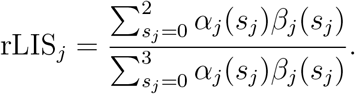

The rejection rule can be written as

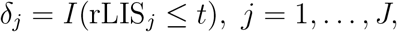

where *I*(*·*) is the indicator function.

We next derive the threshold 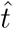 for a pre-specified FDR level *q*. Total number of discoveries and the number of false discoveries are 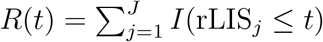 and 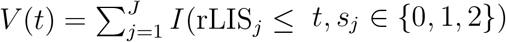, respectively. We have

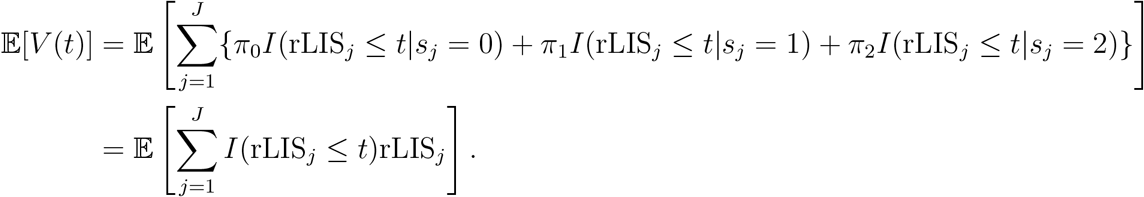

Let rLIS_(1)_ *≤* rLIS_(2)_ *≤ … ≤* rLIS_(*J*)_ be the order statistics and *H*_(1)_, …, *H*_(*J*)_ be the corresponding hypotheses. If *k* hypotheses are rejected, the number of false discoveries can be estimated by

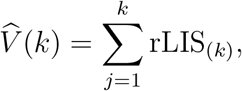

and the FDR can be estimated by 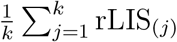. We shall use the following step-up procedure to control the FDR at level *q* (Sun and Cai, 2009).

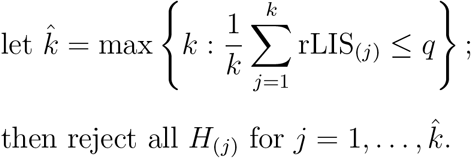

We provide an estimation of ***λ*** in the next section.

#### 4.2.2 Data-driven testing procedure

To estimate the unknown parameters and functions in ***λ***, we first define two posterior probabilities *γ*_*j*_(*s*_*j*_) = 𝕡_***λ***_(*s*_*j*_ | ***p***_1_, ***p***_2_) and *ξ*_*j*_(*s*_*j*_, *s*_*j*+1_) = 𝕡_***λ***_(*s*_*j*_, *s*_*j*+1_ | ***p***_1_, ***p***_2_). By the definition, 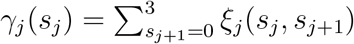. They can be obtained from the forward and backward probabilities

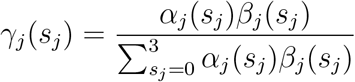

and

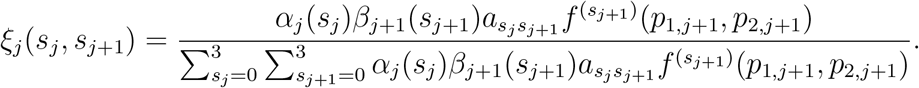

The likelihood function of the complete data (***p***_1_, ***p***_2_, ***s***) is given by

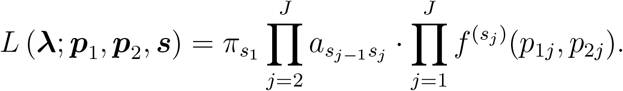

We develop a non-parametric EM algorithm (Dempster et al., 1977) to estimate the unknowns ***λ*** = (***π***, *A, f*_1_, *f*_2_) under the monotone likelihood ratio constraint (3). With an appropriate initialization of the unknowns, ***λ***^(0)^ = (***π***^(0)^, *𝒜* ^(0)^, 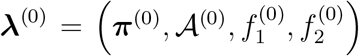), the EM algorithm proceeds by iteratively implementing the following two steps.

**E-step:** Given current 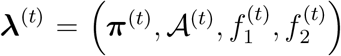, the forward and backward probabilities 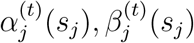 and the posterior probabilities 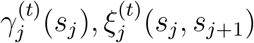 are calculated. The conditional expectation of the log-likelihood function can be written as

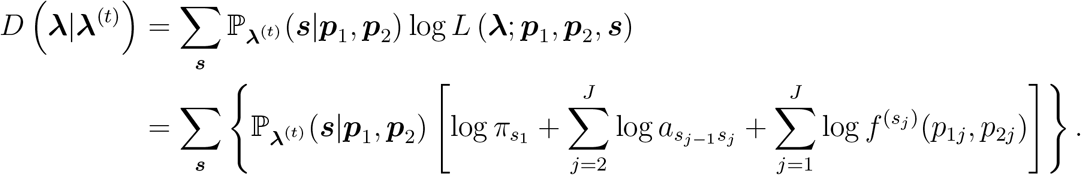

**M-step:** Update ***λ***^(*t*+1)^ by

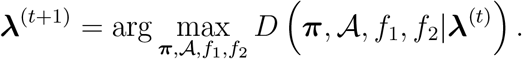

We can update each component alternatingly. By using the Lagrange multiplier, we can calculate ***π***^(*t*+1)^ and *𝒜* ^(*t*+1)^ as

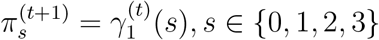

and

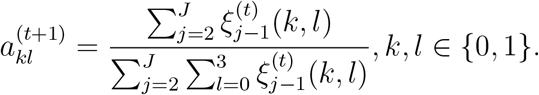

The two functions can be updated by

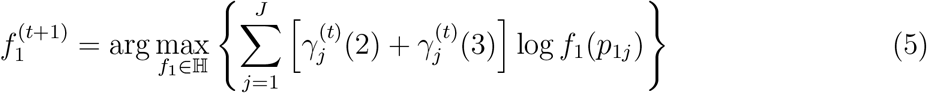

and

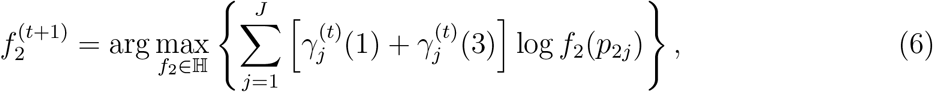

where ℍ is a set of monotonic non-increasing density functions (Sun and Cai, 2007; Cao et al., 2013, 2022). We solve (5) and (6) independently using the non-parametric maximum likelihood estimation implemented with PAVA (Robertson et al., 1988).

The **E-step** and **M-step** are conducted iteratively until convergence. Detailed derivations of the algorithm are presented in the Supplemental Note A. With the estimate 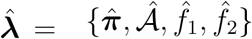, we can calculate the test statistics 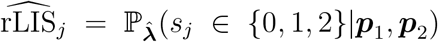. Let 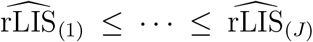 be the order statistics of 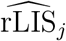, and denote *H*_(1)_, …, *H*_(*J*)_ as the corresponding *H*_0*j*_. The data-driven testing procedure works as follows.

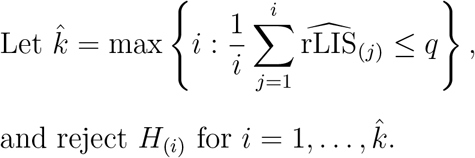

### 4.3 Realistic simulation design

In simulation II, we perform realistic simulations to show the rankings of SNPs using rLIS in two GWASs, where LD structures are derived from real data. Based on the CEU genotype data and FIN genotype data from the Genetic European Variation in Disease project (Lappalainen et al., 2013), we filter out SNPs with the same genotypes in all samples and obtain genotypes of 16, 764 SNPs in both studies. We specify 5 causal SNPs in each study, 4 of which are the same in the two studies. Two of the 4 causal SNPs are close (separated by five SNPs), and the other SNPs are selected randomly. Then in each study, for the *i*th subject (*i* = 1, …, 78 in the CEU study and *i* = 1, …, 89 in the FIN study), we generate continuous phenotypes using the linear regression model

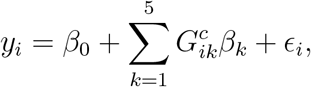

where *β*_0_ is the intercept term, 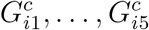 are the genotypes of the *i*th subject for the 5 causal SNPs, *β*_1_, …, *β*_5_ are regression coefficients, and *ϵ*_*i*_ is an error term generated from *N* (0, 1), a standard normal distribution. The intercept term and the regression coefficients of causal SNPs *β*_*k*_, *k* = 0, 1, …, 5, follow *N* (0, *σ*^2^), a normal distribution with mean 0 and standard deviation *σ*. We set *σ* = 0.6 such that the SNP heritability 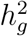, similar in spirit to the *R*^2^ in linear regression models, is centered between 0.2 and 0.3. *p*-values are obtained by a marginal regression of each SNP on the phenotype. We repeat the above process 100 times to get *p*-values of 1, 676, 400 SNPs.

### 4.4 Computation time

We compare the computation time of different methods. All methods are implemented in R, in which STAREG and ReAD use Rcpp to speed up the computation. All computations are carried out in an Intel(R) Core(TM) i7-9750H 2.6GHz CPU with 64 GB RAM laptop. Table 3 summarizes the results. We observe that all methods are quick to compute. The additional time that ReAD takes in simulation studies is negligible in practice.

**Table 3:** Computation time (in seconds) of different methods.

| Dataset | # of SNPs | MaxP | STAREG | ReAD |
| --- | --- | --- | --- | --- |
| Simulation I | 10,000 | 0.0033 | 0.0165 | 0.1871 |
| Simulation II | 1,676,400 | 0.1897 | 3.7237 | 35.159 |
| Asthma | 6,222,195 | 0.7317 | 169.82 | 105.47 |
| UC | 6,232,147 | 0.7428 | 9.5248 | 83.974 |

## Data Availability

All data produced in the present study are available upon reasonable request to the authors.

## Notes

### Competing Interest Statement

The authors have declared no competing interest.

### Funding Statement

This study did not receive any funding.

### Author Declarations

The study used (or will use) ONLY openly available human data that were originally located at the Trans-National Asthma Genetic Consortium (https://www.ebi.ac.uk/gwas/downloads/summary-statistics),the International Inflammatory Bowel Disease Genetics Consortium (http://www.ibdgenetics.org/), and the UK Biobank (http://www.ukbiobank.ac.uk/).

