## Supplementary for "A powerful replicability analysis of genome-wide association studies"

### A Estimating unknown parameters and functions

The forward and backward probabilities $\alpha_j(s_j) = \mathbb{P}_{\boldsymbol{\lambda}}\left((p_{1t}, p_{2t})_{t=1}^{j}, s_j\right)$ and $\beta_j(s_j) = \mathbb{P}_{\boldsymbol{\lambda}}\left((p_{1t}, p_{2t})_{t=j+1}^{J} \mid s_j\right)$ can be calculated recursively using the forward-backward procedure (Baum et al. 1970). Specifically, we initialize $\alpha_1(s_1) = \pi_{s_1} f^{(s_1)}(p_{11}, p_{21})$ and $\beta_J(s_J) = 1$. We have

$$\begin{aligned}
\alpha_{j+1}(s_{j+1}) &= \mathbb{P}_{\boldsymbol{\lambda}}\left((p_{1t}, p_{2t})_{t=1}^{j+1}, s_{j+1}\right) \\
&= \sum_{s_j=0}^{3} \mathbb{P}_{\boldsymbol{\lambda}}((p_{1t}, p_{2t})_{t=1}^{j}, (p_{1,j+1}, p_{2,j+1})|s_j, s_{j+1})\mathbb{P}(s_{j+1}|s_j)\mathbb{P}(s_j) \\
&= \sum_{s_j=0}^{3} \mathbb{P}_{\boldsymbol{\lambda}}\left((p_{1t}, p_{2t})_{t=1}^{j}|s_j\right)\mathbb{P}(s_j)\mathbb{P}_{\boldsymbol{\lambda}}(p_{1,j+1}, p_{2,j+1}|s_{j+1})a_{s_j s_{j+1}} \\
&= \sum_{s_j=0}^{3} \alpha_j(s_j)a_{s_j s_{j+1}} f^{(s_{j+1})}(p_{1,j+1}, p_{2,j+1}),
\end{aligned}$$

where we use the Markovian property in the third equation and the definition of $\alpha_j(s_j)$ in the fourth equation. Note that

$$\begin{aligned}
\beta_j(s_j) &= \mathbb{P}_{\boldsymbol{\lambda}}\left((p_{1t}, p_{2t})_{t=j+1}^{J}|s_j\right) \\
&= \sum_{s_{j+1}=0}^{3} \mathbb{P}_{\boldsymbol{\lambda}}\left((p_{1t}, p_{2t})_{t=j+1}^{J}, s_{j+1}|s_j\right) \\
&= \sum_{s_{j+1}=0}^{3} \mathbb{P}_{\boldsymbol{\lambda}}\left((p_{1t}, p_{2t})_{t=j+1}^{J}|s_{j+1}, s_j\right)\mathbb{P}(s_{j+1}|s_j) \\
&= \sum_{s_{j+1}=0}^{3} \mathbb{P}_{\boldsymbol{\lambda}}\left((p_{1t}, p_{2t})_{t=j+1}^{J}|s_{j+1}\right)\mathbb{P}(s_{j+1}|s_j) \\
&= \sum_{s_{j+1}=0}^{3} \mathbb{P}_{\boldsymbol{\lambda}}\left((p_{1t}, p_{2t})_{t=j+2}^{J}|s_{j+1}\right)\mathbb{P}_{\boldsymbol{\lambda}}(p_{1,j+1}, p_{2,j+1}|s_{j+1})\mathbb{P}(s_{j+1}|s_j) \\
&= \sum_{s_{j+1}=0}^{3} \beta_{j+1}(s_{j+1}) f^{(s_{j+1})}(p_{1,j+1}, p_{2,j+1})a_{s_j s_{j+1}},
\end{aligned}$$

where we use the Markovian property in the fourth equation.

The marginal probability density function of the observations $(\boldsymbol{p}_1, \boldsymbol{p}_2)$ at SNP $j$ given $\boldsymbol{\lambda}$ is

$$\begin{aligned}
\mathbb{P}_{\boldsymbol{\lambda}}(\boldsymbol{p}_1, \boldsymbol{p}_2) &= \mathbb{P}_{\boldsymbol{\lambda}}\left((p_{1j}, p_{2j})_{j=1}^{J}\right) \\
&= \sum_{s_j=0}^{3} \mathbb{P}_{\boldsymbol{\lambda}}\left((p_{1j}, p_{2j})_{j=1}^{J}, s_j\right) \\
&= \sum_{s_j=0}^{3} \mathbb{P}_{\boldsymbol{\lambda}}\left((p_{1j}, p_{2j})_{j=1}^{J} | s_j\right) \mathbb{P}(s_j) \\
&= \sum_{s_j=0}^{3} \mathbb{P}_{\boldsymbol{\lambda}}\left((p_{1t}, p_{2t})_{t=1}^{j} | s_j\right) \mathbb{P}(s_j) \mathbb{P}_{\boldsymbol{\lambda}}\left((p_{1t}, p_{2t})_{t=j+1}^{J} | s_j\right) \\
&= \sum_{s_j=0}^{3} \mathbb{P}_{\boldsymbol{\lambda}}\left((p_{1t}, p_{2t})_{t=1}^{j}, s_j\right) \mathbb{P}_{\boldsymbol{\lambda}}\left((p_{1t}, p_{2t})_{t=j+1}^{J} | s_j\right) \\
&= \sum_{s_j=0}^{3} \alpha_j(s_j)\beta_j(s_j).
\end{aligned}$$

The posterior probabilities $\gamma_j(s_j)$ and $\xi_j(s_j, s_{j+1})$ can be obtained by the forward and backward probabilities at SNP $j$. Specifically,

$$\begin{aligned}
\gamma_j(s_j) &= \mathbb{P}_{\boldsymbol{\lambda}}(s_j | \boldsymbol{p}_1, \boldsymbol{p}_2) \\
&= \frac{\mathbb{P}_{\boldsymbol{\lambda}}(s_j, \boldsymbol{p}_1, \boldsymbol{p}_2)}{\mathbb{P}_{\boldsymbol{\lambda}}(\boldsymbol{p}_1, \boldsymbol{p}_2)} \\
&= \frac{\alpha_j(s_j)\beta_j(s_j)}{\sum_{s_j=0}^{3} \alpha_j(s_j)\beta_j(s_j)},
\end{aligned}$$

and

$$\begin{aligned}
\xi_j(s_j, s_{j+1}) &= \mathbb{P}_{\boldsymbol{\lambda}}(s_j, s_{j+1} | \boldsymbol{p}_1, \boldsymbol{p}_2) \\
&= \frac{\mathbb{P}_{\boldsymbol{\lambda}}(s_j, s_{j+1}, \boldsymbol{p}_1, \boldsymbol{p}_2)}{\mathbb{P}_{\boldsymbol{\lambda}}(\boldsymbol{p}_1, \boldsymbol{p}_2)} \\
&= \frac{\alpha_j(s_j)\beta_{j+1}(s_{j+1}) a_{s_j s_{j+1}} f^{(s_{j+1})}(p_{1,j+1}, p_{2,j+1})}{\sum_{s_j=0}^{3} \sum_{s_{j+1}=0}^{3} \alpha_j(s_j)\beta_{j+1}(s_{j+1}) a_{s_j s_{j+1}} f^{(s_{j+1})}(p_{1,j+1}, p_{2,j+1})},
\end{aligned}$$

where the numerator is derived by

$$
\begin{aligned}
&\mathbb{P}_{\boldsymbol{\lambda}}(s_j, s_{j+1}, \boldsymbol{p}_1, \boldsymbol{p}_2)\\
=&\mathbb{P}_{\boldsymbol{\lambda}}\left(s_j, (p_{1t}, p_{2t})_{t=1}^{j}\right)\mathbb{P}_{\boldsymbol{\lambda}}\left(s_{j+1}, (p_{1t}, p_{2t})_{t=j+1}^{J} | s_j\right)\\
=&\alpha_j(s_j)\mathbb{P}_{\boldsymbol{\lambda}}\left((p_{1t}, p_{2t})_{t=j+2}^{J} | s_j, s_{j+1}, p_{1,j+1}, p_{2,j+1}\right)\mathbb{P}_{\boldsymbol{\lambda}}\left(s_{j+1}, p_{1,j+1}, p_{2,j+1} | s_j\right)\\
=&\alpha_j(s_j)\mathbb{P}_{\boldsymbol{\lambda}}\left((p_{1t}, p_{2t})_{t=j+2}^{J} | s_{j+1}\right)\mathbb{P}_{\boldsymbol{\lambda}}\left(s_{j+1}, p_{1,j+1}, p_{2,j+1} | s_j\right)\\
=&\alpha_j(s_j)\beta_{j+1}(s_{j+1})\mathbb{P}_{\boldsymbol{\lambda}}\left(s_{j+1} | s_j\right)\mathbb{P}_{\boldsymbol{\lambda}}\left(p_{1,j+1}, p_{2,j+1} | s_j, s_{j+1}\right)\\
=&\alpha_j(s_j)\beta_{j+1}(s_{j+1})\mathbb{P}_{\boldsymbol{\lambda}}\left(s_{j+1} | s_j\right)\mathbb{P}_{\boldsymbol{\lambda}}\left(p_{1,j+1}, p_{2,j+1} | s_{j+1}\right)\\
=&\alpha_j(s_j)\beta_{j+1}(s_{j+1})a_{s_j, s_{j+1}} f^{(s_{j+1})}(p_{1,j+1}, p_{2,j+1}),
\end{aligned}
$$

3 where we use the Markovian property in the third and fifth equation.

In the **M-step** of the EM algorithm, we update $\boldsymbol{\lambda}^{(t+1)}$ alternatingly by

$$
\boldsymbol{\lambda}^{(t+1)} = \arg\max_{\boldsymbol{\pi}, \mathcal{A}, f_1, f_2} D\left(\boldsymbol{\pi}, \mathcal{A}, f_1, f_2 | \boldsymbol{\lambda}^{(t)}\right).
$$

Recall the definition $\gamma_j(s_j) = \mathbb{P}_{\boldsymbol{\lambda}}(s_j \mid \boldsymbol{p}_1, \boldsymbol{p}_2)$, we first update $\boldsymbol{\pi}^{(t+1)}$ by

$$
\begin{aligned}
\boldsymbol{\pi}^{(t+1)} &= \arg\max_{\boldsymbol{\pi}} \left\{ \sum_{s_1=0}^{3} \sum_{s_2=0}^{3} \cdots \sum_{s_J=0}^{3} (\log \pi_{s_1}) \mathbb{P}_{\boldsymbol{\lambda}^{(t)}}(\boldsymbol{s} | \boldsymbol{p}_1, \boldsymbol{p}_2) \right\}\\
&= \arg\max_{\boldsymbol{\pi}} \left\{ \sum_{s_1=0}^{3} (\log \pi_{s_1}) \sum_{s_2=0}^{3} \cdots \sum_{s_{J-1}=0}^{3} \mathbb{P}_{\boldsymbol{\lambda}^{(t)}}(s_1, \ldots, s_{J-1} | \boldsymbol{p}_1, \boldsymbol{p}_2) \right\}\\
&= \arg\max_{\boldsymbol{\pi}} \left\{ \sum_{s_1=0}^{3} (\log \pi_{s_1}) \mathbb{P}_{\boldsymbol{\lambda}^{(t)}}(s_1 | \boldsymbol{p}_1, \boldsymbol{p}_2) \right\}\\
&= \arg\max_{\boldsymbol{\pi}} \left\{ \sum_{s_1=0}^{3} (\log \pi_{s_1}) \gamma_1^{(t)}(s_1) \right\}, \text{ s.t. } \sum_{s_1=0}^{3} \pi_{s_1} = 1.
\end{aligned}
$$

We use the Lagrange multiplier to solve the maximization. Specifically,

$$L_\pi(\boldsymbol{\pi}, \eta) = \sum_{i=0}^{3} (\log \pi_i) \gamma_1^{(t)}(i) + \eta \left( \sum_{i=0}^{3} \pi_i - 1 \right)$$

By taking a derivative with respect to $\pi_i$, we have

$$\begin{aligned}
\frac{\partial L_\pi(\boldsymbol{\pi}, \eta)}{\partial \pi_i} = \frac{\gamma_1^{(t)}(i)}{\pi_i} + \eta = 0 \Rightarrow \\
\eta \pi_i = -\gamma_1^{(t)}(i) \Rightarrow \\
\eta \sum_{i=0}^{3} \pi_i = -\sum_{i=0}^{3} \gamma_1^{(t)}(i) \Rightarrow \\
\eta = -1,
\end{aligned}$$

where we use the property that $\sum_{i=0}^{3} \pi_i = \sum_{i=0}^{3} \gamma_1^{(t)}(i) = 1$. Consequently, we have

$$\pi_i^{(t+1)} = \gamma_1^{(t)}(i).$$

Next, we update $\mathcal{A}^{(t+1)}$ by

$$\begin{aligned}
\mathcal{A}^{(t+1)} &= \arg\max_{\mathcal{A}} \left\{ \sum_{\boldsymbol{s}} \mathbb{P}_{\boldsymbol{\lambda}^{(t)}}(\boldsymbol{s} | \boldsymbol{p}_1, \boldsymbol{p}_2) \sum_{j=2}^{J} \log a_{s_{j-1} s_j} \right\} \\
&= \arg\max_{\mathcal{A}} \left\{ \sum_{\boldsymbol{s}} \mathbb{P}_{\boldsymbol{\lambda}^{(t)}}(\boldsymbol{s} | \boldsymbol{p}_1, \boldsymbol{p}_2) \log a_{s_1 s_2} + \cdots + \sum_{\boldsymbol{s}} \mathbb{P}_{\boldsymbol{\lambda}^{(t)}}(\boldsymbol{s} | \boldsymbol{p}_1, \boldsymbol{p}_2) \log a_{s_{J-1} s_J} \right\}.
\end{aligned}$$

The target function is the summation of $J-1$ terms with the same form. For the first term, we have

$$\begin{aligned}
&\sum_{\boldsymbol{s}} \mathbb{P}_{\boldsymbol{\lambda}^{(t)}}(\boldsymbol{s}|\boldsymbol{p}_1, \boldsymbol{p}_2) \log a_{s_1 s_2} \\
=&\sum_{s_1} \sum_{s_2} \cdots \sum_{s_J} \mathbb{P}_{\boldsymbol{\lambda}^{(t)}}(s_1, \ldots, s_J|\boldsymbol{p}_1, \boldsymbol{p}_2) \log a_{s_1 s_2} \\
=&\sum_{s_1} \sum_{s_2} \mathbb{P}_{\boldsymbol{\lambda}^{(t)}}(s_1, s_2|\boldsymbol{p}_1, \boldsymbol{p}_2) \log a_{s_1 s_2}.
\end{aligned}$$

Hence the problem can be represented as

$$\begin{aligned}
\mathcal{A}^{(t+1)} &= \arg\max_{\mathcal{A}} \left\{ \sum_{k=0}^{3} \sum_{l=0}^{3} \log a_{kl} \sum_{j=2}^{J} \mathbb{P}_{\boldsymbol{\lambda}^{(t)}}(s_{j-1} = k, s_j = l|\boldsymbol{p}_1, \boldsymbol{p}_2) \right\}, \\
\text{s.t. } & \sum_{l=0}^{3} a_{kl} = 1, k \in \{0, 1, 2, 3\}.
\end{aligned}$$

Using the Lagrange multiplier, we have the objective function

$$L_A(\mathcal{A}, \eta) = \sum_{k=0}^{3} \sum_{l=0}^{3} \log a_{kl} \sum_{j=2}^{J} \mathbb{P}_{\boldsymbol{\lambda}^{(t)}}(s_{j-1} = k, s_j = l|\boldsymbol{p}_1, \boldsymbol{p}_2) + \sum_{k=0}^{3} \eta_k \left( \sum_{l=0}^{3} a_{kl} - 1 \right).$$

Taking a derivative with respect to $a_{kl}$, we have

$$\begin{aligned}
\frac{\partial L_a(\mathcal{A}, \eta)}{\partial a_{kl}} &= \frac{1}{a_{kl}} \sum_{j=2}^{J} \mathbb{P}_{\boldsymbol{\lambda}^{(t)}}(s_{j-1} = k, s_j = l|\boldsymbol{p}_1, \boldsymbol{p}_2) + \eta_k = 0 \Rightarrow \\
a_{kl} &= -\frac{\sum_{j=2}^{J} \mathbb{P}_{\boldsymbol{\lambda}^{(t)}}(s_{j-1} = k, s_j = l|\boldsymbol{p}_1, \boldsymbol{p}_2)}{\eta_k}. \qquad \text{(S1)}
\end{aligned}$$

As $\sum_{l=0}^{3} a_{kl} = 1$, we have

$$\begin{aligned}
\sum_{l=0}^{3} a_{kl} &= -\frac{\sum_{l=0}^{3} \sum_{j=2}^{J} \mathbb{P}_{\boldsymbol{\lambda}^{(t)}}(s_{j-1} = k, s_j = l|\boldsymbol{p}_1, \boldsymbol{p}_2)}{\eta_k} = 1 \Rightarrow \\
\eta_k &= -\sum_{j=2}^{J} \mathbb{P}_{\boldsymbol{\lambda}^{(t)}}(s_{j-1} = k|\boldsymbol{p}_1, \boldsymbol{p}_2).
\end{aligned}$$

Recall the definition $\xi_j(s_j, s_{j+1}) = \mathbb{P}_{\boldsymbol{\lambda}}(s_j, s_{j+1} \mid \boldsymbol{p}_1, \boldsymbol{p}_2)$ and take into account Equation (S1), we obtain $a_{kl}^{(t+1)}$ by plugging in $\eta_k$,

$$\begin{aligned} a_{kl}^{(t+1)} &= \frac{\sum_{j=2}^{J} \mathbb{P}_{\boldsymbol{\lambda}^{(t)}}(s_{j-1} = k, s_j = l|\boldsymbol{p}_1, \boldsymbol{p}_2)}{\sum_{j=2}^{J} \mathbb{P}_{\boldsymbol{\lambda}^{(t)}}(s_{j-1} = k|\boldsymbol{p}_1, \boldsymbol{p}_2)} \\ &= \frac{\sum_{j=2}^{J} \xi_{j-1}^{(t)}(k, l)}{\sum_{j=2}^{J} \sum_{l=0}^{3} \xi_{j-1}^{(t)}(k, l)}. \end{aligned}$$

Finally, we update $f_1^{(t+1)}$ and $f_2^{(t+1)}$. Note that

$$\begin{aligned} \left(f_1^{(t+1)}, f_2^{(t+1)}\right) &= \arg\max_{f_1, f_2} \left\{ \sum_{\boldsymbol{s}} \mathbb{P}_{\boldsymbol{\lambda}^{(t)}}(\boldsymbol{s}|\boldsymbol{p}_1, \boldsymbol{p}_2) \sum_{j=1}^{J} \log f^{(s_j)}(p_{1j}, p_{2j}) \right\} \\ &= \arg\max_{f_1, f_2} \Bigg\{ \sum_{\boldsymbol{s}} \left[\mathbb{P}_{\boldsymbol{\lambda}^{(t)}}(\boldsymbol{s}|\boldsymbol{p}_1, \boldsymbol{p}_2) \log f^{(s_1)}(p_{11}, p_{21})\right] + \cdots + \\ &\qquad \sum_{\boldsymbol{s}} \left[\mathbb{P}_{\boldsymbol{\lambda}^{(t)}}(\boldsymbol{s}|\boldsymbol{p}_1, \boldsymbol{p}_2) \log f^{(s_J)}(p_{1J}, p_{2J})\right] \Bigg\}. \end{aligned}$$

Recall the definition $\gamma_j(s_j) = \mathbb{P}_{\boldsymbol{\lambda}}(s_j \mid \boldsymbol{p}_1, \boldsymbol{p}_2)$, for the first term, we have

$$\begin{aligned} &\sum_{\boldsymbol{s}} \left[\mathbb{P}_{\boldsymbol{\lambda}^{(t)}}(\boldsymbol{s}|\boldsymbol{p}_1, \boldsymbol{p}_2) \log f^{(s_j)}(p_{1j}, p_{2j})\right] \\ &= \sum_{s_j=0}^{3} \log f^{(s_j)}(p_{1j}, p_{2j}) \sum_{s_1=0}^{3} \cdots \sum_{s_{j-1}=0}^{3} \sum_{s_{j+1}=0}^{3} \cdots \sum_{s_J=0}^{3} \mathbb{P}_{\boldsymbol{\lambda}^{(t)}}(s_1, \ldots, s_J|\boldsymbol{p}_1, \boldsymbol{p}_2) \\ &= \sum_{s_j=0}^{3} \log f^{(s_j)}(p_{1j}, p_{2j}) \mathbb{P}_{\boldsymbol{\lambda}^{(t)}}(s_j|\boldsymbol{p}_1, \boldsymbol{p}_2) \\ &= \sum_{s_j=0}^{3} \gamma_j^{(t)}(s_j) \log f^{(s_j)}(p_{1j}, p_{2j}). \end{aligned}$$

The target function to maximize becomes

$$
\begin{aligned}
L_f(f_1, f_2) =& \sum_{j=1}^{J} \sum_{s_j=0}^{3} \gamma_j^{(t)}(s_j) \log f^{(s_j)}(p_{1j}, p_{2j}) \\
=& \sum_{j=1}^{J} \Big[ \gamma_j^{(t)}(0) \log f_0(p_{1j}) f_0(p_{2j}) + \gamma_j^{(t)}(1) \log f_0(p_{1j}) f_2(p_{2j}) + \\
& \gamma_j^{(t)}(2) \log f_1(p_{1j}) f_0(p_{2j}) + \gamma_j^{(t)}(3) \log f_1(p_{1j}) f_2(p_{2j}) \Big].
\end{aligned}
$$

Then $f_1$ and $f_2$ can be updated by

$$
f_1^{(t+1)} = \arg\max_{f_1 \in \mathbb{H}} \left\{ \sum_{j=1}^{J} \left[ \gamma_j^{(t)}(2) + \gamma_j^{(t)}(3) \right] \log f_1(p_{1j}) \right\}
$$

and

$$
f_2^{(t+1)} = \arg\max_{f_2 \in \mathbb{H}} \left\{ \sum_{j=1}^{J} \left[ \gamma_j^{(t)}(1) + \gamma_j^{(t)}(3) \right] \log f_2(p_{2j}) \right\}.
$$

We provide specific steps as follows. Denote $Q_{1j}^{(t)} = \gamma_j^{(t)}(2) + \gamma_j^{(t)}(3)$ and $Q_{2j}^{(t)} = \gamma_j^{(t)}(1) + \gamma_j^{(t)}(3)$, where $j = 1, \dots, J$. Let $0 = p_{1(0)} \leq p_{1(1)} \leq \cdots \leq p_{1(J)}$ be the order statistics of $\boldsymbol{p}_1$ and denote $Q_{1(j)}^{(t)}$ as the corresponding $Q_{1j}^{(t)}$. Let $0 = p_{2(0)} \leq p_{2(1)} \leq \cdots \leq p_{2(J)}$ be the order statistics of $\boldsymbol{p}_2$ and denote $Q_{2(j)}^{(t)}$ as the corresponding $Q_{2j}^{(t)}$. Define $y_{1j} = f_1\left(p_{1(j)}\right)$ and $y_{2j} = f_2\left(p_{2(j)}\right)$, then we can write (2) as

$$
\begin{aligned}
f_1^{(t+1)} &= \arg\max_{y_{1j} \in \mathcal{M}_1} \left\{ \sum_{j=1}^{J} Q_{1(j)}^{(t)} \log y_{1j} \right\}, \text{ subject to } \sum_{j=1}^{J} y_{1j}(p_{1(j)} - p_{1(j-1)}) = 1, \text{ and} \\
f_2^{(t+1)} &= \arg\max_{y_{2j} \in \mathcal{M}_2} \left\{ \sum_{j=1}^{J} Q_{2(j)}^{(t)} \log y_{2j} \right\}, \text{ subject to } \sum_{j=1}^{J} y_{2j}(p_{2(j)} - p_{2(j-1)}) = 1,
\end{aligned}
$$

where $\mathcal{M}_1 = \{(y_{11}, \dots, y_{1J}) : y_{11} \geq \cdots \geq y_{1J} \geq 0\}$ and $\mathcal{M}_2 = \{(y_{21}, \dots, y_{2J}) : y_{21} \geq \cdots \geq y_{2J} \geq 0\}$.

Using the Lagrangian multiplier, the objective functions we want to maximize are

$$\sum_{j=1}^{J} Q_{1(j)}^{(t)} \log y_{1j} + \eta_1 \left\{ \sum_{j=1}^{J} y_{1j}(p_{1(j)} - p_{1(j-1)}) - 1 \right\}, \text{ and}$$
$$\sum_{j=1}^{J} Q_{2(j)}^{(t)} \log y_{2j} + \eta_2 \left\{ \sum_{j=1}^{J} y_{2j}(p_{2(j)} - p_{2(j-1)}) - 1 \right\}.$$

Taking derivatives with respect to $y_{1j}, \eta_1$ and $y_{2j}, \eta_2$, respectively, we have

$$\hat{\eta}_1 = -\sum_{j=1}^{J} Q_{1(j)}^{(t)}, \ \tilde{y}_{1j} = \frac{Q_{1(j)}^{(t)}}{Q_1^{(t)}\big(p_{1(j)} - p_{1(j-1)}\big)}, \text{ and}$$
$$\hat{\eta}_2 = -\sum_{j=1}^{J} Q_{2(j)}^{(t)}, \ \tilde{y}_{2j} = \frac{Q_{2(j)}^{(t)}}{Q_2^{(t)}\big(p_{2(j)} - p_{2(j-1)}\big)},$$

6 where $Q_1^{(t)} = \sum_{j=1}^{J} Q_{1j}^{(t)}$ and $Q_2^{(t)} = \sum_{j=1}^{J} Q_{2j}^{(t)}$.

To incorporate the monotone constraints on $y_{1j}$ and $y_{2j}$, we minimize

$$\sum_{j=1}^{J} \left\{ -Q_{1(j)}^{(t)} \log y_{1j} + Q_1^{(t)}(p_{1(j)} - p_{1(j-1)}) y_{1j} \right\} = \sum_{j=1}^{J} Q_{1(j)}^{(t)} \left\{ -\log y_{1j} - \frac{-Q_1^{(t)}\big(p_{1(j)} - p_{1(j-1)}\big)}{Q_{1(j)}^{(t)}} y_{1j} \right\}$$

subject to $y_{11} \geq \cdots \geq y_{1J}$, and minimize

$$\sum_{j=1}^{J} \left\{ -Q_{2(j)}^{(t)} \log y_{2j} + Q_2^{(t)}(p_{2(j)} - p_{2(j-1)}) y_{2j} \right\} = \sum_{j=1}^{J} Q_{2(j)}^{(t)} \left\{ -\log y_{2j} - \frac{-Q_2^{(t)}\big(p_{2(j)} - p_{2(j-1)}\big)}{Q_{2(j)}^{(t)}} y_{2j} \right\}$$

7 subject to $y_{21} \geq \cdots \geq y_{2J}$.

Let

$$(\hat{u}_{11}, \ldots, \hat{u}_{1J}) = \arg \min_{u_{11}, \ldots, u_{1J}} \sum_{j=1}^{J} Q_{1(j)}^{(t)} \left( u_{1j} - \frac{-Q_1^{(t)}\big(p_{1(j)} - p_{1(j-1)}\big)}{Q_{1(j)}^{(t)}} \right)^2$$

subject to $u_{11} \geq u_{12} \geq \cdots \geq u_{1J}$, and

$$(\hat{u}_{21}, \ldots, \hat{u}_{2J}) = \arg \min_{u_{21}, \ldots, u_{2J}} \sum_{j=1}^{J} Q_{2(j)}^{(t)} \left( u_{2j} - \frac{-Q_2^{(t)} \left( p_{2(j)} - p_{2(j-1)} \right)}{Q_{2(j)}^{(t)}} \right)^2$$

subject to $u_{21} \geq u_{22} \geq \cdots \geq u_{2J}$. The solutions take the max-min form

$$\hat{u}_{1j} = \max_{b \geq j} \min_{a \leq j} \frac{-Q_1^{(t)} \sum_{k=a}^{b} \left( p_{1(k)} - p_{1(k-1)} \right)}{\sum_{k=a}^{b} Q_{1(k)}^{(t)}},$$
$$\hat{u}_{2j} = \max_{b \geq j} \min_{a \leq j} \frac{-Q_2^{(t)} \sum_{k=a}^{b} \left( p_{2(k)} - p_{2(k-1)} \right)}{\sum_{k=a}^{b} Q_{2(k)}^{(t)}},$$

which can be obtained by PAVA (Busing 2022). Our final estimates are given by $\hat{y}_{1j} = -\frac{1}{\hat{u}_{1j}}$ and $\hat{y}_{2j} = -\frac{1}{\hat{u}_{2j}}$ for $j = 1, \ldots, J$ according to Theorem 3.1 of Barlow & Brunk (1972).

### B Methods comparison

To evaluate the performance of ReAD. We compare the FDR and power of ReAD with several replicability analysis methods, including STAREG, *ad hoc* BH, MaxP (Benjamini et al. 2009), JUMP (Lyu et al. 2023), radjust (Bogomolov & Heller 2018) and MaRR (Philtron et al. 2018). We review the details of these methods as follows.

#### B.1 The STAREG method

Let $\tau_j = (\theta_{1j}, \theta_{2j}),\ j = 1, \ldots, J$ denote the inferred association status of SNPs across two studies. Then $\tau_j \in \{(0,0), (0,1), (1,0), (1,1)\}$ with $\mathbb{P}(\tau_j = (k,l)) = \xi_{kl}$ for $k, l = 0, 1$ and

$\sum_{k,l} \xi_{kl} = 1$. Assume a mixture model for $p$-values in the two studies. Specifically,

$$p_{1j} \mid \theta_{1j} \sim (1 - \theta_{1j}) f_0 + \theta_{1j} f_1,$$
$$p_{2j} \mid \theta_{2j} \sim (1 - \theta_{2j}) f_0 + \theta_{2j} f_2, \quad j = 1, \ldots, J,$$

where $f_0$ is the density function of $p$-values under the null, $f_1$ and $f_2$ denote the non-null density functions for study 1 and study 2, respectively. Then the local false discovery rate (Lfdr) is defined as the posterior probability of being replicability null given data. We have

$$\begin{aligned}\mathrm{Lfdr}_j(p_{1j}, p_{2j}) &:= 1 - \mathbb{P}(\theta_{1j} = \theta_{2j} = 1 \mid p_{1j}, p_{2j}) \\ &= \frac{\xi_{00} f_0(p_{1j}) f_0(p_{2j}) + \xi_{01} f_0(p_{1j}) f_2(p_{2j}) + \xi_{10} f_1(p_{1j}) f_0(p_{2j})}{\xi_{00} f_0(p_{1j}) f_0(p_{2j}) + \xi_{01} f_0(p_{1j}) f_2(p_{2j}) + \xi_{10} f_1(p_{1j}) f_0(p_{2j}) + \xi_{11} f_1(p_{1j}) f_2(p_{2j})}.\end{aligned}$$

Assume the monotone likelihood ratio condition (Sun & Cai 2007, Cao et al. 2013, 2022):

$$f_1(x)/f_0(x) \text{ and } f_2(x)/f_0(x) \text{ are non-increasing in } x. \tag{S2}$$

We have that $\mathrm{Lfdr}_j$ is monotonically non-decreasing in $(p_{1j}, p_{2j})$. The rejection rule based on $\mathrm{Lfdr}_j$ to test the replicability null is $\delta_j = I\{\mathrm{Lfdr}_j \leq \lambda\}$, where $\lambda$ is a threshold to be determined. We write the total number of discoveries as $R(\lambda) = \sum_{j=1}^{J} I\{\mathrm{Lfdr}_j \leq \lambda\}$, and the number of false discoveries as $V(\lambda) = \sum_{j=1}^{J} I\{\mathrm{Lfdr}_j \leq \lambda\}(1 - \theta_{1j}\theta_{2j})$. In the oracle case that we know $(\xi_{00}, \xi_{01}, \xi_{10}, \xi_{11}, f_1, f_2)$, define

$$\lambda_J = \sup\left\{\lambda \in [0,1] : \frac{\sum_{j=1}^{J} \mathrm{Lfdr}_j I\{\mathrm{Lfdr}_j \leq \lambda\}}{\sum_{j=1}^{J} I\{\mathrm{Lfdr}_j \leq \lambda\}} \leq q\right\}.$$

16 Reject $H_{0j}$ if $\mathrm{Lfdr}_j \leq \lambda_J$. Then the FDR is asymptotically controlled at level $q$.

Assume $f_0$ follows a standard uniform distribution. Let $\boldsymbol{p}_1 = \{p_{1j}\}_{j=1}^{J}$ and $\boldsymbol{p}_2 = \{p_{2j}\}_{j=1}^{J}$ denote $p$-values from study 1 and study 2, respectively. Denote $\boldsymbol{\theta}_1 = \{\theta_{1j}\}_{j=1}^{J}$ and $\boldsymbol{\theta}_2 =$

$\{\theta_{2j}\}_{j=1}^J$. The unknown parameters and functions are estimated by maximizing the following log-likelihood function

$$
\begin{aligned}
l(\boldsymbol{p}_1, \boldsymbol{p}_2, \boldsymbol{\theta}_1, \boldsymbol{\theta}_2) = \sum_{j=1}^{J} \big[ & \log\{(1-\theta_{1j})f_0(p_{1j}) + \theta_{1j} f_1(p_{1j})\} + \log\{(1-\theta_{2j})f_0(p_{2j}) + \theta_{2j} f_2(p_{2j})\} \\
& + \theta_{1j}(1-\theta_{2j}) \log \xi_{10} + (1-\theta_{1j})\theta_{2j} \log \xi_{01} + (1-\theta_{1j})(1-\theta_{2j}) \log \xi_{00} \\
& + \theta_{1j}\theta_{2j} \log \xi_{11} \big],
\end{aligned}
$$

where $\boldsymbol{\theta}_1$ and $\boldsymbol{\theta}_2$ are latent variables. For scalable computation, we utilize EM algorithm (Dempster et al. 1977) in combination of pool-adjacent-violator-algorithm (PAVA) (Robertson et al. 1988) to efficiently estimate the unknowns $(\xi_{00}, \xi_{01}, \xi_{10}, \xi_{11}, f_1, f_2)$ incorporating the monotonic constraint (S2) for $f_1$ and $f_2$. With the estimates $(\hat{\xi}_{00}, \hat{\xi}_{01}, \hat{\xi}_{10}, \hat{\xi}_{11}, \hat{f}_1, \hat{f}_2)$, we obtain the estimated Lfdr as follows.

$$
\widehat{\text{Lfdr}}_j = \frac{\hat{\xi}_{00} f_0(p_{1j}) f_0(p_{2j}) + \hat{\xi}_{01} f_0(p_{1j}) \hat{f}_2(p_{2j}) + \hat{\xi}_{10} \hat{f}_1(p_{1j}) f_0(p_{2j})}{\hat{\xi}_{00} f_0(p_{1j}) f_0(p_{2j}) + \hat{\xi}_{01} f_0(p_{1j}) \hat{f}_2(p_{2j}) + \hat{\xi}_{10} \hat{f}_1(p_{1j}) f_0(p_{2j}) + \hat{\xi}_{11} \hat{f}_1(p_{1j}) \hat{f}_2(p_{2j})}.
$$

An estimate of $\lambda_J$ is

$$
\hat{\lambda}_J = \sup \left\{ \lambda \in [0,1] : \frac{\sum_{j=1}^J \widehat{\text{Lfdr}}_j I\{\widehat{\text{Lfdr}}_j \le \lambda\}}{\sum_{j=1}^J I\{\widehat{\text{Lfdr}}_j \le \lambda\}} \le q \right\}.
$$

The replicability null hypothesis $H_{0j}$ is rejected if $\widehat{\text{Lfdr}}_j \le \hat{\lambda}_J$. This is equivalent to the step-up procedure (Sun & Cai 2007): let $\widehat{\text{Lfdr}}_{(1)} \le \ldots \le \widehat{\text{Lfdr}}_{(J)}$ be the order statistics of $\{\widehat{\text{Lfdr}}_j\}_{j=1}^J$ and denote by $H_{(1)}, \ldots, H_{(J)}$ the corresponding ordered hypotheses, the procedure works as follows.

$$\text{Find} \quad \hat{k} := \max\left\{k \in [1,m] : \frac{1}{k}\sum_{j=1}^{k} \widehat{\text{Lfdr}}_{(j)} \leq \alpha \right\}, \text{ and}$$

$$\text{reject } H_{(j)}, \quad j = 1, \ldots, \hat{k}.$$

#### B.2 The *ad hoc* BH method

BH (Benjamini & Hochberg 1995) is the most popular multiple testing procedure that conservatively controls the FDR for $J$ independent or positively dependent tests. In study $i$, $i = 1, 2$, the BH procedure proceeds as follows.

- *Step 1.* Let $p_{i(1)} \leq p_{i(2)} \leq \cdots \leq p_{i(J)}$ be the ordered $p$-values, and denote by $H^i_{(j)}$ the corresponding hypothesis;
- *Step 2.* Find the largest $k$ such that $p_{i(k)} \leq \frac{k}{J}q$, i.e., $\hat{k} = \max\{1 \leq k \leq J : p_{i(k)} \leq \frac{k}{J}q\}$, and $\hat{k} = 0$ if the set is empty;
- *Step 3.* Reject $H^i_{(j)}, j = 1, \ldots, \hat{k}$.

The *ad hoc* BH method for replicability analysis identifies SNPs rejected by both studies as replicable SNPs.

#### B.3 The MaxP method

Define the maximum of $p$-values as

$$p_j^{\max} = \max\{p_{1j}, p_{2j}\}, j = 1, \ldots, J.$$

$p_j^{\max}$ follows a super-uniform distribution under the replicability null. The MaxP method directly applies BH (Benjamini & Hochberg 1995) to $p_j^{\max}, j = 1, \ldots, J$ for FDR control.

#### B.4 The JUMP method

The JUMP method (Lyu et al. 2023) works on the maximum of $p$-values across two studies. Define

$$p_j^{\max} = \max\{p_{1j}, p_{2j}\}, j = 1, \ldots, J.$$

Let $\tau_j = (\theta_{1j}, \theta_{2j}),\ j = 1, \ldots, J$ denote the inferred association status of SNPs across two studies. Then $\tau_j \in \{(0,0), (0,1), (1,0), (1,1)\}$ with $\mathbb{P}(\tau_j = (k,l)) = \xi_{kl}$ for $k, l = 0, 1$ and $\sum_{k,l} \xi_{kl} = 1$. It can be shown that

$$\begin{aligned}
&\mathbb{P}\left(p_j^{\max} \le t \mid H_{0j} \text{ is true}\right) \\
=&\frac{\xi_{00}\mathbb{P}(p_j^{\max} \le t \mid \tau_j = (0,0))}{\xi_{00} + \xi_{01} + \xi_{10}} + \frac{\xi_{01}\mathbb{P}(p_j^{\max} \le t \mid \tau_j = (0,1))}{\xi_{00} + \xi_{01} + \xi_{10}} + \frac{\xi_{10}\mathbb{P}(p_j^{\max} \le t \mid \tau_j = (1,0))}{\xi_{00} + \xi_{01} + \xi_{10}} \\
\le&\frac{\xi_{00}t^2 + (\xi_{01} + \xi_{10})t}{\xi_{00} + \xi_{01} + \xi_{10}} \le t,
\end{aligned}$$

which means that $p_j^{\max}$ follows a super-uniform distribution under the replicability null. Denote

$$G(t) = \frac{\xi_{00}t^2 + (\xi_{01} + \xi_{10})t}{\xi_{00} + \xi_{01} + \xi_{10}}.$$

For a given threshold $t \in (0,1)$, a conservative estimate of the FDR is obtained by

$$\text{FDR}^*(t) = \frac{J(\xi_{00} + \xi_{01} + \xi_{10})G(t)}{\sum_{j=1}^{J} I\{p_j^{\max} \le t\} \vee 1}.$$

Following Storey (2002), Storey et al. (2004), the proportion of null hypotheses in study $i$ can be estimated by

$$\hat{\pi}_0^{(i)}(\lambda_i) = \frac{\sum_{j=1}^{J} I\{p_{ij} \ge \lambda_i\}}{J(1 - \lambda_i)}, \quad i = 1, 2.$$

Similarly, $\xi_{00}$ is estimated by

$$\hat{\xi}_{00}(\lambda_3) = \frac{\sum_{j=1}^{J} I\{p_{1j} \geq \lambda_3, p_{2j} \geq \lambda_3\}}{J(1-\lambda_3)^2},$$

where $\lambda_1, \lambda_2$ and $\lambda_3$ are tuning parameters that can be selected by using the smoothing method provided in Storey & Tibshirani (2003). Then we have

$$\hat{\xi}_{01} = \hat{\pi}_0^{(1)} - \hat{\xi}_{00}, \quad \hat{\xi}_{10} = \hat{\pi}_0^{(2)} - \hat{\xi}_{00}.$$

With these estimates, we have a plug-in estimate of FDR,

$$\widehat{\mathrm{FDR}}^*(t) = \frac{J(\hat{\xi}_{00}t^2 + \hat{\xi}_{01}t + \hat{\xi}_{10}t)}{\sum_{j=1}^{J} I\{p_j^{\max} \leq t\} \vee 1}.$$

The JUMP method works as follows.

- *Step 1.* Let $p_{(1)}^{\max} \leq \cdots \leq p_{(J)}^{\max}$ be the ordered maximum of $p$-values and denote by $H_{(j)}$ the corresponding hypothesis;

- *Step 2.* Find the largest $k$ such that the estimated FDR is controlled, i.e.,

$$\hat{k} = \max\{1 \leq k \leq J : \widehat{\mathrm{FDR}}^*(p_{(k)}^{\max}) \leq q\};$$

- *Step 3.* Reject $H_{(j)}$, $j = 1, \ldots, \hat{k}$.

#### B.5 The radjust procedure

The radjust procedure (Bogomolov & Heller 2018) works as follows,

- *Step 1.* For a pre-specified FDR level $q$, compute

$$R = \max\left[r : \sum_{j \in \mathcal{S}_1 \cap \mathcal{S}_2} I\left\{(p_{1j}, p_{2j}) \leq \left(\frac{rq}{2|\mathcal{S}_2|}, \frac{rq}{2|\mathcal{S}_1|}\right)\right\} = r\right],$$

  where $\mathcal{S}_i$ is the set of features pre-selected in study $i$ for $i = 1, 2$. By default, it selects features with $p$-values less than or equal to $q/2$.

- *Step 2.* Reject features with indices in the set

$$\mathcal{R} = \left\{j : (p_{1j}, p_{2j}) \leq \left(\frac{Rq}{2|\mathcal{S}_2|}, \frac{Rq}{2|S_1|}\right), j \in \mathcal{S}_1 \cap \mathcal{S}_2\right\}.$$

In this paper, we implement an adaptive version of the radjust procedure Bogomolov & Heller (2018) in the simulations, which first estimates the fractions of true null hypotheses among the pre-selected features. The fractions in the two studies are estimated as follows.

$$\hat{\pi}_0^{(1)} = \frac{1 + \sum_{j \in \mathcal{S}_{2,q}} I(p_{1j} > q)}{|\mathcal{S}_{2,q}|(1-q)}, \quad \hat{\pi}_0^{(2)} = \frac{1 + \sum_{j \in \mathcal{S}_{1,q}} I(p_{2j} > q)}{|\mathcal{S}_{1,q}|(1-q)}, \tag{S3}$$

where $\mathcal{S}_{i,q} = \mathcal{S}_i \cap \{1 \leq j \leq J : p_{ij} \leq q\},\ i = 1, 2$. The adaptive procedure with a nominal FDR level $q$ works as follows.

- *Step 1.* Compute $\hat{\pi}_0^{(1)}$ and $\hat{\pi}_0^{(2)}$ using (S3). Let

$$R = \max\left[r : \sum_{j \in \mathcal{S}_{1,q} \cap \mathcal{S}_{2,q}} I\left\{(p_{1j}, p_{2j}) \leq \left(\frac{rq}{2|\mathcal{S}_{2,q}|\hat{\pi}_0^{(1)}}, \frac{rq}{2|\mathcal{S}_{1,q}|\hat{\pi}_0^{(2)}}\right)\right\} = r\right],$$

- *Step 2.* Reject features with indices in the set

$$\mathcal{R} = \left\{j : (p_{1j}, p_{2j}) \leq \left(\frac{Rq}{2|\mathcal{S}_{2,q}|\hat{\pi}_0^{(1)}}, \frac{Rq}{2|S_{1,q}|\hat{\pi}_0^{(2)}}\right), j \in \mathcal{S}_{1,q} \cap \mathcal{S}_{2,q}\right\}.$$

#### B.6 The MaRR procedure

The MaRR procedure (Philtron et al. 2018) uses the maximum rank of each feature. The null hypothesis is that $H_{0j}$ : $p_{1j}$ and $p_{2j}$ are irreplicable. Denote $(R_{1j}, R_{2j})$ as the ranks of $(p_{1j}, p_{2j}), j = 1, \dots, J$ within each study. Define

$$M_j = \max\{R_{1j}, R_{2j}\}, i = 1, \dots, J.$$

Let $\pi_1$ denote the proportion of replicable features. Under the assumptions:
(I1) if gene $g$ is replicable and gene $h$ is irreplicable

$$R_{1g} < R_{1h}, \quad R_{2g} < R_{2h};$$

(I2) the correlation between the ranks of replicable features is non-negative;
(I3) the two ranks of irreplicable genes are independent.
Under these assumptions, irreplicable ranks $R_{1j}$ and $R_{2j}$ are uniformly distributed between $\lfloor J\pi_1 \rfloor + 1$ and $J$. Denote the conditional null survival function of $M_j/J$ as

$$\begin{aligned}
S_{J,\pi_1}(x) &= P(M_j/J > x \mid \text{ gene } j \text{ is irreplicable}) \\
=& 1 - P\left(R_{1j}/J \leq x, R_{2j}/J \leq x \mid \text{ gene } j \text{ is irreplicable}\right) \\
=& 1 - \prod_{i=1}^{2} P\left(R_{ij}/J \leq x \mid \text{ gene } j \text{ is irreplicable}\right) \\
=& \begin{cases} 1, & x < \pi_1, \\ 1 - \frac{(j_x - i_{\pi_1})^2}{(J - i_{\pi_1})^2}, & \pi_1 \leq x \leq 1, \end{cases}
\end{aligned}$$

where $j_x = \lfloor Jx \rfloor$ and $i_{\pi_1} = \lfloor J\pi_1 \rfloor$. The limiting conditional survival function of $M_j/J$ under the null is

$$S_{J,\pi_1}(x) \to S_{\pi_1}(x) = \begin{cases} 1 & x < \pi_1 \\ 1 - \frac{(x-\pi_1)^2}{(1-\pi_1)^2} & \pi_1 \le x \le 1 \\ 0 & 1 < x. \end{cases}$$

The empirical survival function can be estimated by $\hat{S}_J(x) = \frac{1}{J}\sum_{i=1}^{J} I(M_j/J \ge x)$, $x \in (0,1)$. By strong law of large numbers and the Bayesian formula, we have

$$\begin{aligned} \hat{S}_J(x) \to & P(M_j/J \ge x) \\ = & (1-\pi_1)P(M_j/J \ge x \mid \text{ gene } j \text{ is irreplicable}) + \pi_1 \times 0 \\ = & (1-\pi_1)S_{\pi_1}(x) \text{ for } x \in (\pi_1, 1). \end{aligned}$$

If we estimate $\pi_1$ by $j/J$, we can define the mean square error (MSE) as follows.

$$\text{MSE}(j/J) = (J-j)^{-1} \sum_{k=j}^{J} \Big( \hat{S}_J(k/J) - (1-j/J)S_{j/J}(k/J) \Big)^2 .$$

$\hat{k}$ is chosen to minimize the MSE in the range between 0 and $\lfloor 0.9J \rfloor$.

$$\hat{k} = \mathop{\arg\min}_{j=0,1,\ldots,\lfloor 0.9J \rfloor} \{\text{MSE}(j/J)\} .$$

Thus $\hat{k}/J$ serves as a good estimate of $\pi_1$. To control the FDR at level $q$, MaRR generates the rejection threshold as follows.

$$\text{Define } \hat{N} = \max_{\hat{k} < j \le n} \left\{ j : m\widehat{\text{FDR}}(j) = \frac{(j-\hat{k})^2}{Q(j)(J-\hat{k})} \le q \right\},$$

where $Q(j) = \sum_{k=1}^{J} I(M_k \leq j)$. Reject features associated with $M_j \leq \hat{N}$. Philtron et al. (2018) relaxes assumption (I1) to (R1): $P(R_{1g} < R_{1h}) > 1/2$ and $P(R_{2g} < R_{2h}) > 1/2$, which is more plausible in practice.

### C Simulation details

In simulation I, the hidden states of SNPs, $(s_j)_{j=1}^{J} \in \{0, 1, 2, 3\}$, are generated from a four-state Markov chain, where the initial probabilities of the four states are $\boldsymbol{\pi}^0 = (0.9, 0.025, 0.025, 0.05)$, and the transition matrix is

$$\mathcal{A} = \begin{pmatrix} a_{00} & (1-a_{00})/3 & (1-a_{00})/3 & (1-a_{00})/3 \\ (1-a_{11})/3 & a_{11} & (1-a_{11})/3 & (1-a_{11})/3 \\ (1-a_{22})/3 & (1-a_{22})/3 & a_{22} & (1-a_{22})/3 \\ (1-a_{33})/3 & (1-a_{33})/3 & (1-a_{33})/3 & a_{33} \end{pmatrix}.$$

$\theta_{ij}, i = 1, 2, j = 1, \ldots, J$, can be obtained from $(s_j)_{j=1}^{J}$. Denote $N(\mu, \sigma^2)$ the normal distribution with mean $\mu$ and variance $\sigma^2$. We generate $z$-statistics from a mixture model $X_{ij} \mid \theta_{ij} \sim (1-\theta_{ij})N(0,1) + \theta_{ij}N(\mu_i, 1)$ for $i = 1, 2$ and $j = 1, \ldots, J$, where $\mu_i$ represents the signal strength of study $i$. Corresponding one-sided $p$-values are calculated by $p_{ij} = 1 - \Phi(X_{ij})$, where $\Phi(\cdot)$ is the cumulative distribution function of the standard normal distribution $N(0, 1)$.

In addition to *ad hoc* BH, MaxP, and STAREG, we also compare the performance of ReAD to more replicability analysis methods, including JUMP (Lyu et al. 2023), radjust (Bogomolov & Heller 2018) and MaRR (Philtron et al. 2018). Let $J = 10,000, \mu_1 = 2$ and $a_{00} = a_{11} = a_{22}$ in all simulations. We vary $a_{00}, a_{33}$ and $\mu_2$ to evaluate the FDR and power of different methods in different simulation settings. Empirical FDR and power are calculated from 100 runs for each setting. In Fig. S1 (left: FDR; right: power), each row corresponds to a different $a_{00}$, and each column corresponds to a different $a_{33}$. In each panel, we set $\mu_2$

to be 1.5, 2, or 3. At FDR level 0.05, we see that the *ad hoc* BH fails to control the FDR in many settings. STAREG has a slight FDR inflation in some settings. The other methods control the FDR at the target level across all settings, in which MaxP and radjust are overly conservative, and rLIS shows substantial power gain. In addition, the power of all methods increases with increased signal strength ($\mu_2$).

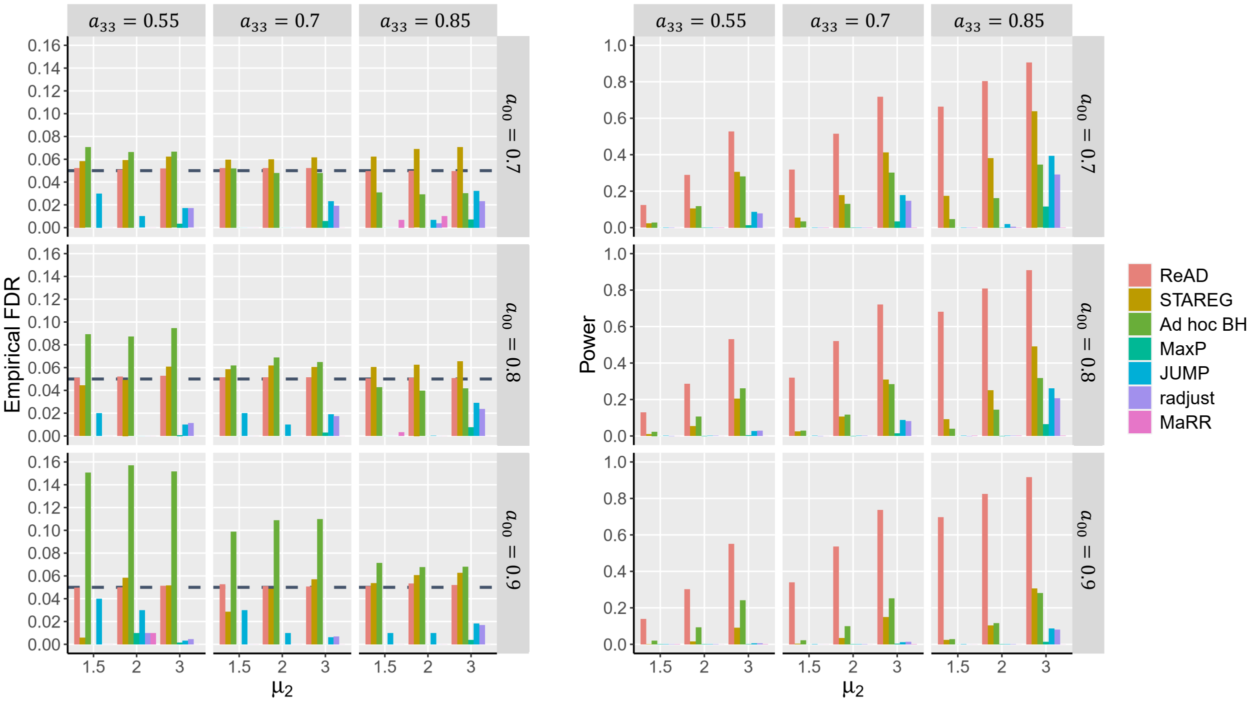


Figure S1: FDR control and power comparison of different methods
